# A quantitative systems pharmacology workflow towards optimal design and biomarker stratification of atopic dermatitis clinical trials

**DOI:** 10.1101/2023.07.01.23292105

**Authors:** Natacha Go, Simon Arsène, Igor Faddeenkov, Théo Galland, Shiny Martis B., Diane Lefaudeux, Yishu Wang, Loic Etheve, Evgueni Jacob, Claudio Monteiro, Jim Bosley, Caterina Sansone, Christian Pasquali, Lorenz Lehr, Alexander Kulesza

## Abstract

**Background:** The development of atopic dermatitis (AD) drugs is confronted by many disease phenotypes and trial design options, which are hard to explore experimentally.

**Objective:** Optimize AD trial design using simulations.

**Methods:** We constructed a quantitative systems pharmacology (QSP) model of AD and standard of care (SoC) treatments and generated a phenotypically diverse virtual population whose parameter distribution is derived from known relationships between AD biomarkers and disease severity and b) calibrated using disease severity evolution under SoC regimens.

**Results:** We applied this workflow to the immunomodulator OM-85, currently being investigated for its potential use in AD, and calibrated investigational treatment model with the efficacy profile of an existing trial (thereby enriching it with plausible marker levels and dynamics). We assessed the sensitivity of trial outcomes to trial protocol and found that for this particular example, a) the choice of endpoint is more important than the choice of dosing-regimen and b) patient selection by model-based responder enrichment could increase the expected effect size. A global sensitivity analysis reveals that only a limited subset of baseline biomarkers is needed to predict the drug response of the full virtual population

**Conclusion:** This AD QSP workflow built around knowledge of marker-severity relationships as well as SoC efficacy can be tailored to specific development cases so as to optimize several trial protocol parameters and biomarker-stratificaiton and therefore holds promise to become a powerful model-informed drug development tool.

**Key Messages:**

- Disease and treatment models can quantify pre-existing knowledge about complex immune diseases such as atopic dermatitis and drug’s efficacy data under one common umbrella.
- Embedding QSP models into trial simulation setup can give insight into clinical trial optimization.
- Complex QSP models can help with patient selection and biomarker identification.

**Capsule Summary:** This study shows the relevance of QSP model and computer simulations in assisting clinical development in the field of atopic dermatitis by assessing the impact of trial protocol on treatment effect and guiding biomarker programs.

## 1 Introduction

Atopic dermatitis (AD) is a chronic inflammatory condition of the skin characterized by recurrent eczematous lesions and intense itch which can profoundly impair quality of life^1–6^. The pathophysiology of AD is complex and involves local and systemic immune dysregulation, genetic susceptibility, environmental factors and microbiome effects^7;8^. AD is not a homogeneous disease encompasses a variety of endotypes and phenotypes in the different age, ethnic, etc. groups (see the work cited in Facheris et al. ^9^). The road to novel therapeutics^10^ is tortuous, due a combination of two factors:

a. a partial understanding of the molecular and cellular mechanisms driving disease severity and treatment effect in individual patients^11;12^, making it difficult to translate preclinical results into the clinic and even more so to estimate the quantitative effect in a given population.
b. the complex clinical management of AD^13;14^ which opens many up many potential choices in trial design such as rules for rescue treatment or definition of induction or washout periods^15^.

Despite advancement in our understanding of epidemiology, biomarkers, endotypes, prevention, and comorbidities^16^ as well as *in vitro*, *in vivo* and *in silico* approaches developed for the investigation of human AD pathogenesis^17^, the precise relationships between intrinsic immunological features, extrinsic factors, disease severity and heterogeneous treatment effect remains to be established. *In silico* approaches based on mechanistic models can be powerful tools to explore trial design options^18;19^ in a context, such as AD, where high heterogeneity makes that, in addition to questions on protocol and dosing-regimen, the strategies involving patient selection, involving for example local or systemic biomarkers, are of utmost importance^20^. There are several existing mathematical models for AD^17^, notably highlighting the importance of temporality in skin barrier function, immune responses, and impact of environmental stressors^21^ as well as the identification of effective biologic drugs combination for single-drug non-responders^22^. These studies have demonstrated that a systems modeling approach can help resolving mechanistic questions in AD drug development. Still, the question of how to quantitatively translate these insights into model-informed clinical trial design remains open. Within model-informed drug development, QSP models are being increasingly considered, both for internal and regulatory decision making^23^. QSP - including within a trial simulation paradigm - has produced numerous published examples and regulatory grade evidence across disease areas where protocols and in particular the dosing-regimen aspects have been informed^23;24^, but the use of QSP for informing predictive biomarker programs have been mostly limited to generating plausible dynamics of important biomarkers in different tissues and did not yield quantitative and actionable information on clinical applications such as e.g. a biomarker identification trial. This gap motivated us to combine a QSP modeling approach for an investigational treatment in AD with a trial simulation strategy aiming at both, quantifying the relationship between trial design options and the expected efficacy profiles while taking into account the patients’ immune profiles and thereby addressing biomarker related trial design choices.

In this work, we consider the investigational treatment OM-85 as a use-case. The oral immunomodulator OM-85 is a bacterial lysate which modulates the immune system notably by restoring the Th1/Th2 balance^25^ and is thus being investigated for various atopic conditions^26^. OM-85 is approved in several countries for the prevention of respiratory tract infections^27^ and has shown some promising results in pediatric AD as add-on treatment^28^. It is now being tested for early moderate AD in an ongoing trial (NCT05222516) where the immune heterogeneity of the included population can represent a challenge. We developed a novel mechanistic model of skin immune dysregulation in AD including the mechanisms of action of emollients and topical corticosteroids (TCS), combined with a previously developed model of the administration, pharmacokinetics, and mechanism of action of OM-85 (originally applied in combination with a multiscale respiratory tract infection disease model^29^). We calibrated this QSP model with quantitative relationships in skin biomarkers^30–35^, clinical data for standard of care (SoC)^22;36–39^ and OM-85 treatment efficacy^28^. As a plausible application example, we used this QSP model to run trial simulations for virtual AD populations in order to obtain both patient- and trial protocol-specific predictions of drug efficacy profiles. We found that trial efficacy is expected to be insensitive to the exact dosing regimen but can display sensitivity with respect to choice of severity endpoint (relative vs absolute), which thus appears to be an option for further trial optimization. We then show how a dimensionality reduction approach can be used to inform a biomarker-based stratification strategy: global sensitivity analysis identifies a reduced number of baseline biomarkers in the virtual population, that reproduce the reference data well, and thus can be used to select patients based on their predicted treatment effect. We finally show that in the chosen use case, both protocol design and patient selection with results of the model do show optimization potential and therefore may ultimately support decisions about clinical (predictive) biomarker programs.

## 2 Methods

### 2.1 QSP workflow: combining systems immunology and trial simulations

Our QSP workflow (modeling and simulation approach) is based on a knowledge and mechanism-driven mathematical model focusing on the immunology of AD, built on the back of a system of ordinary differential equations (ODEs) and used in a setup to simulate clinical trials (also referred to as a quantitative disease-drug-trial model^40^ or *in silico* clinical trials). A virtual population approach captures between-patient variability, where model parameters are described by statistical (co)distributions rather than scalar values.

### 2.2 QSP model description

The QSP model (Fig. 1) can be broken down into three components: a) the disease model describing the skin barrier integrity and the immune system and linking baseline biomarkers levels to AD severity scores, b) the SoC treatment models accounting for the administration and mechanisms of action of emollients and TCS, and c) the investigational treatment model describing the administration, pharmacokinetics, and mechanism of action of OM-85. The disease model focuses on selected quantitative aspects of skin immune dysregulation in patients with AD while not taking into account pathogenesis or natural evolution of AD. This allowed us to consider that, without treatment, the disease state of a given patient with a given AD severity can be characterized by stationary dynamics of the biological entities. Under treatment however, the stationary immune dynamics of the disease model (*i.e.* at baseline) are perturbed and the AD severity evolves. The calibration of the QSP model was performed following the core approach for complex disease-treatment-trial models with heterogeneous data as described in Palgen et al. ^41^.

**Figure 1.**
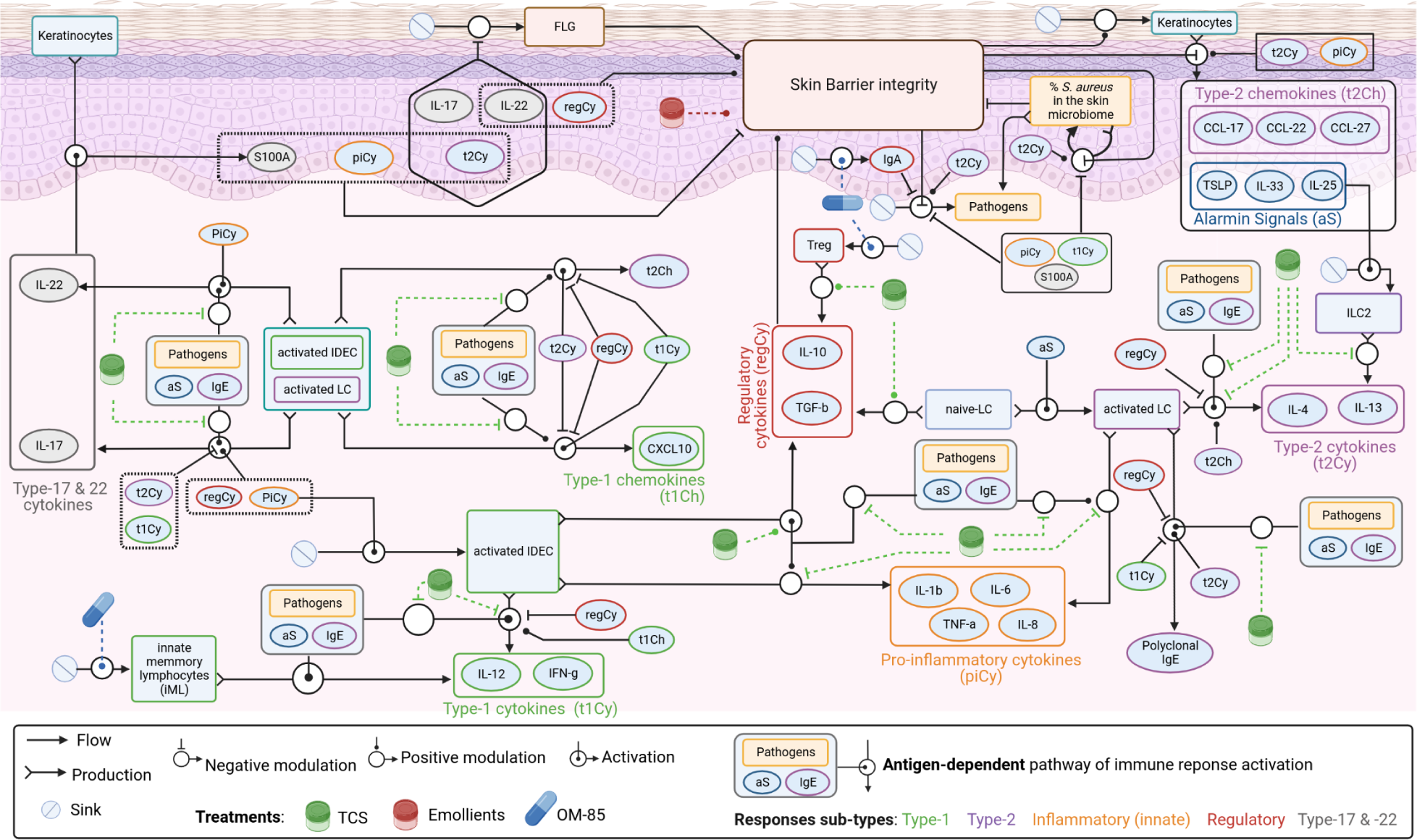
QSP model schematic. Exhaustive representation of the interplay between the skin barrier and the skin immune system as formalized in the AD disease model, mechanisms of actions of SoC treatments (TCS and emollients) and the investigational treatment OM-85.

#### 2.2.1 Atopic dermatitis disease model

The disease model covers the main mechanisms at the cellular scale involved in the modulation of the skin barrier integrity and immune dysregulation in response to environmental stressors. In particular, it captures the mechanisms driving the dynamics of the major biomarkers of AD lesional skin including filaggrin, *S. aureus* in the skin microbiome, and major soluble immune biomarkers (mainly cytokines and chemokines characterizing the type-1, 2, 17, 22, and regulatory responses). While for typical pharmacometrics analyses, the principle of parsimony guides the selection of the model structure^42^, our mechanistic model is complex and combines a heterogenous set of information from different sources. Its architecture consists of a system of ODEs representing the dynamics of 21 variables and involves 139 parameters (Fig. 1, Supplementary Information S1.1).

The main mechanisms included in the disease model can be summarized as follows. The skin barrier integrity is modulated by filaggrin levels^43^, harmful *S. aureus* colonization^6;44^, and skin immune dysregulation^6;44–49^. In return, conjointly with immune dysregulation, skin barrier integrity modulates the level of pathogen infiltration^50–52^ and determines the production of alarmins (TSLP, IL-25, IL-33). Alarmins activate the innate immune response, notably Langerhans cells (LC) and inflammatory epidermal dendritic cells (IDEC),^47;52–54^ both in antigen-independent and -dependent pathways (via alarmins or specific IgE, respectively)^55;56^. Both LC and IDEC synthesize innate pro-inflammatory factors (IL-1β, IL-6, TNF-α, IL-8)^53;56^ as well as type-17/22 immunity proteins (IL-17, IL-22, and S100A)^43–45;48;57^. Activated LCs – in context of AD^53^ – induce the polarization of the adaptive response towards type-2 (represented by IL-4, IL-13 and IgE) through the production of type-2 chemokines (CCL-17, CCL-27 and CCL-22), while IDEC polarize it towards type-1 (represented by IL-12 and IFN-γ) through the production of type-1 chemokines (CXCL-10)^56^. The interconnected feedback loops of the type-1 vs type-2 response converge to an overall type-2 skewing in AD, which in return is mixed with growing type-1 response with increasing AD severity^53;56;57^. In AD context, regulatory cytokines (IL-10 and TGF-β) are produced by regulatory T cells (Treg) and dendritic cells (naive LC and activated IDEC with high level of FcεRI on their cell surface)^47;53;56;58^ and limit the innate inflammation as well as type-1 and type-2 responses^44;58;59^. AD severity (evaluated with SCORAD^60^ and EASI^61^) is phenomenologically linked to the level of skin barrier integrity and pathogen infiltration^22^.

We make the fundamental assumption that, without treatment, the disease model represents the skin immune system at equilibrium (Supplementary Information S1.2.1). By computing and applying the mathematical conditions for equilibrium, we are able to derive a large number of parameters (reducing the set of free parameters from 113 to 23, Supplementary Information S1.2.1, Supplementary Information Table S7). We can therefore inform the steady-state values of the thirteen biomarkers included in the model as a function of AD severity for individual patients based on a collection of datasets from the literature (Supplementary Information S1.2.2, Supplementary Information Table 4). Note that the entire dynamic system is still underinformed meaning that there are still parameters for which several values satisfy the conditions for equilibrium. For this reason it is important to also constrain the dynamical behavior upon perturbation of the equilibrium, i.e. under treatment (where target engagement and pathway modulation will drive this system out of equilibrium) as described in what follows.

#### 2.2.2 Standard of care treatment model

The SoC treatment comprises a combination of emollients and different TCS (Fig. 1). The application of emollient ameliorates the skin barrier integrity^3;62;63^ while TCS inhibit type-1, type-2 and pro-inflammatory responses and amplify the regulatory response by modulating respective cytokines^64–68^. We disregarded topical drug pharmacokinetics by considering a standardized bioavailable concentration throughout a day of treatment regardless of duration of application, dosage and intra-daily frequency. We also neglected the potential systemic effects given the very low systemic absorption of modern TCS^69;70^. We calibrated the SoC treatments against an aggregated dataset of 5 studies accounting for various combinations and potencies (Supplementary Information S1.2.3, Supplementary Information Table S6). The set of calibrated parameters include those driving SoC mechanisms of action but also those controlling AD pathophysiology left undetermined after applying equilibrium constraints (Supplementary Information Table S7). Finally, we used the placebo arm (only emollients + TCS) reported in our reference dataset for the investigational treatment (Bodemer et al. ^28^) to assess the robustness of model prediction for SoC treatments.

#### 2.2.3 Investigational treatment (OM-85) model

For OM-85 administration and effect, we re-used the model reported in Arsène et al. ^29^ where, as described in the context of respiratory tract infections prophylaxis, the oral administration of OM-85 triggers the activation and proliferation in the intestinal Peyers Patches of reprogrammed type-1 innate memory like cells^71–73^, regulatory T-cells^74–76^ and polyclonal IgA producing plasmablasts^77;78^, which further disseminate via the systemic circulation into the inflamed lung tissues, due to the presence of chemokines and homing receptors. In the context of AD, the fundamental hypothesis is that that similar cells infiltrate the inflamed skin tissues (the gut-skin axis as a central mechanistic hypothesis), which is backed up notably by many reports of co-expression of gut, skin and lung tissue homing markers^52;58;79;80^ and is consistent with the reported effect of OM-85 in AD^28^. To take this hypothesis into account, the model reported in Arsène et al. ^29^ is supplemented with a layer describing skin-homing of activated immune effectors (Fig. 1, Supplementary Information Figure S1).

### 2.3 Analyses

#### 2.3.1 Treatment effect

The treatment effect can be quantified *in silico* by the individual absolute benefit (AB, see Boissel et al. ^81^) which is defined as the difference of SCORAD at 6 months for a specific virtual patient with placebo vs when treated (in our idealized clinical setting each virtual patient can be its own control).

#### 2.3.2 Average treatment effect

The average treatment effect is the group equivalent of treatment effect, defined as the difference of the mean severity score at 6 months in the placebo vs treatment arm (AB, see e.g. Bodemer et al. ^28^). We obtain confidence intervals around predicted values by using a bootstrapped approach (using 100 samples, each with 85 virtual patients for the placebo and for the treatment arms). For the sensitivity assessment of the trial results with respect to the choice of main endpoint, we used four alternative definitions for the average treatment effect, either based on SCORAD, EASI, relative SCORAD or relative EASI. For relative SCORAD or EASI, the difference in severity scores at 6 months is normalized by the mean severity score at 6 months in the placebo arm.

#### 2.3.3 Power and sample size

We compute empirical statistical power through bootstrapping^82^ using Students t-test (alpha risk = 0.05) for various sample sizes. This allows us to estimate the required sample size to reach a statistical power of 0.80(Supplementary Information S.1.4).

#### 2.3.4 Recruitment effort

We define recruitment effort as the total number of patients to be screened given selection criterion: we compute it by dividing the estimated sample size for the corresponding selection criteria by the percentage of virtual patients matching the selection criteria in the reference virtual population, for which the distribution of patient characteristics is assumed to be realistic.

## 3 Results

### 3.1 Reproduction of a placebo-controlled clinical trial

In order to quantitatively align our QSP model with a specific clinical development case, we adapted a simulation and analysis protocol to represent an existing trial (Bodemer et al. ^28^ [NCT05222516]) and used the reported efficacy profiles to calibrate the remaining undetermined parameters (controlling the effect of OM-85). This simulation protocol corresponds to a placebo-controlled parallel two-arm AD trial for a virtual population of pediatric subjects treated with OM-85 for 9 months and assessing AD severity (SCORAD) during the study (Table 1). In order to match the longitudinal efficacy data for OM-85, we calibrated the three parameters controlling the effects of immune effectors activated by OM-85 against the average evolution of disease severity in the treatment arm (all the other model parameters being already calibrated: see SoC treatment and Arsène et al. ^29^). As illustrated in Fig. 2, average value and variability of disease severity evolution in absolute and in relative values, and value at 6 months (Fig. 2 A-B, C-D, E-F respectively) are well reproduced, for both placebo and treatment arms.

**Figure 2.**
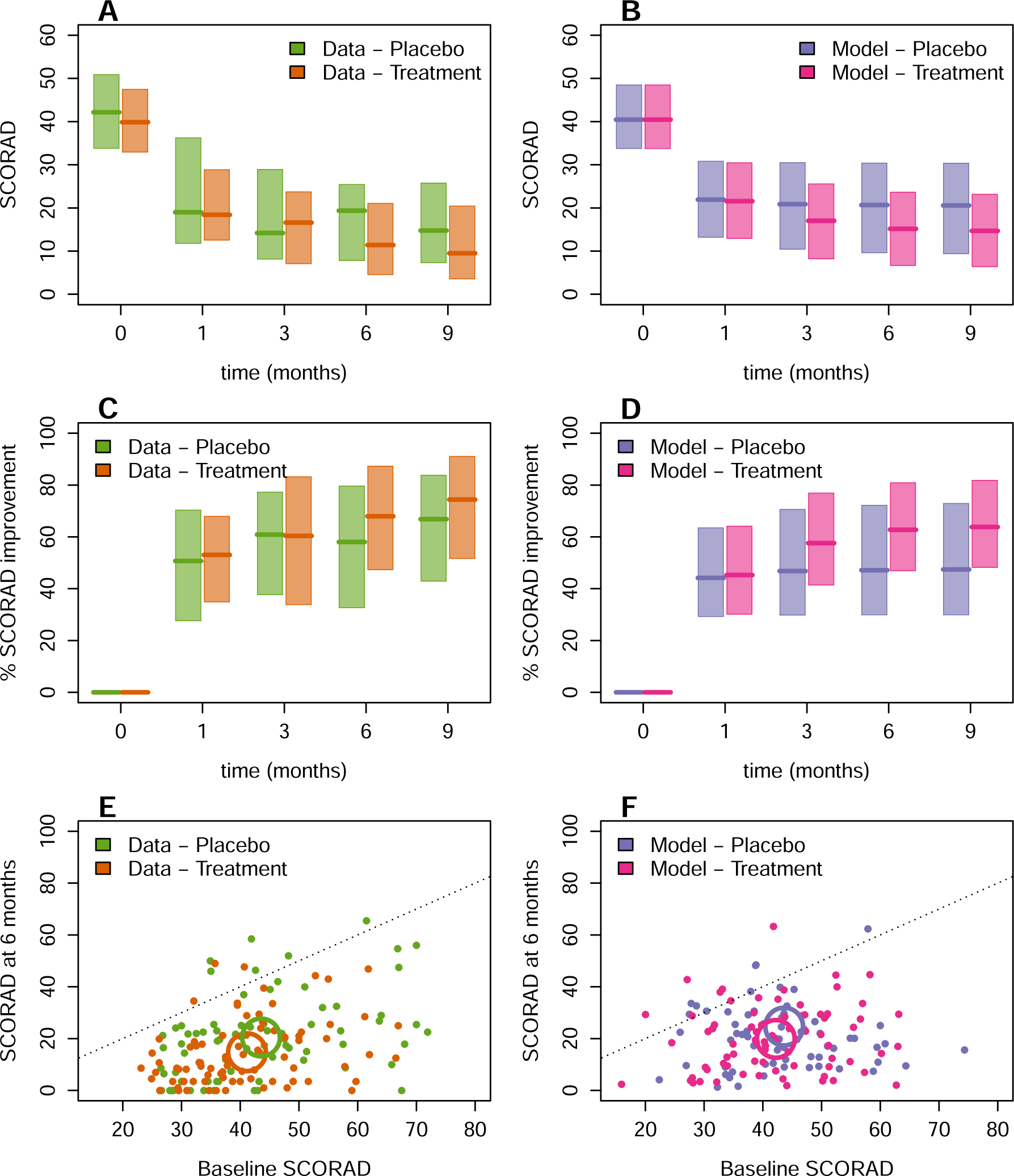
Reproduction of a randomized placebo-controlled clinical trial. A – B: Evolution of AD severity (SCORAD, median: line, quartiles: box) of treatment vs placebo arms from the real trial (Bodemer et al. ^28^) (A) and virtual population (B) at observation time points. C – D. Evolution of percentage improvement of AD severity (SCORAD, median: line, quartiles: box) of treatment vs placebo arms from the real trial (Bodemer et al. ^28^) (C) and virtual population (D) at observation time points. E – F: Individual (points) and average (circles) treatment effect represented by the SCORAD at 6 months compared with baseline SCORAD from the real trial (Bodemer et al. ^28^) (E) and randomly sampled virtual patients matching the baseline SCORAD for placebo and treatment arms (F).

**Table 1.** Simulation protocol for in silico clinical trials mimicking the randomized reference clinical trial in Bodemer et al.^28^ [NCT05222516]. Virtual population (2000 patients / arm), administration regimen, arms, and measurements. * Calibrated.

|  |  |  |  |  |  |
| --- | --- | --- | --- | --- | --- |
| Virtual population | Age | $\ln(\text{Age}) \sim \mathcal{N}(0.3, 1.8) \text{ \& } 0.6 \leq \text{Age} \leq 8$ | | | |
| | Gender | $\sim \mathcal{B}(0.5)$ | | | |
| | SCORAD | $\ln(\text{SCORAD}) \sim \mathcal{N}(3.7, 0.27) \text{ \& } 15 \leq \text{SCORAD} \leq 95$ | | | |
| | Biomarkers | function*(SCORAD) + $\mathcal{N}(\text{mean}^*, \text{sd}^*)$ | | | |
| Administration regimen |  | Week 0 – 1 | Week 1 – 2 | Week 2 – 3 | Week 4 – 36 |
|  | Emollients | Every 1 day |  |  |  |
|  | TCS | Every 1 day | Every 2 days | Every 3 days | Every 3 days (*) |
|  | OM-85 | Every 1 day |  |  |  |
| Arms | Placebo | Emollients + TCS |  |  |  |
|  | Treatment | Emollients + TCS + OM-85 |  |  |  |
| Measurements |  | Baseline (day 0) |  | Follow-up: month 1, 3, 6, 9 |  |
|  | Data<sup>28</sup> | SCORAD, flares |  |  |  |
|  | Model | SCORAD, EASI, biomarkers |  | SCORAD, EASI |  |

In particular, SCORAD at 6 months for data vs model are Q1: 7.7 vs 10.2 median: 19.4 vs 21 Q3: 25.5 vs 31. The average calibration error in median SCORAD for the placebo arm is 4.1 (range: 1.7 7.1), and 3.2 (range: 0.6 5.8) for the treatment arm. These results show that a predicted impact of protocol changes to SCORAD higher than 5 points can be considered to exceed the efficacy prediction error of our QSP model.

### 3.2 Assessment of trial results sensitivity to the trial protocol

Trial protocols are complex and contain a high number of degrees of freedom. As such, their impact on efficacy are typically difficult to assess empirically. To address this difficulty, we intend to show that *in silico* trials based on our QSP model can allow clinicians to systematically test variations in trial protocol and conclude on possible trial outcomes. We therefore explored potential design alternatives for the trial reproduced in Section 3.1 (see also Table 1) in order to identify the trial protocol parameters to which the average treatment effect is most sensitive. We report the average treatment effect and sample size estimated for given power in Fig. 3. Overall, the average treatment effect shows a clear sensitivity to variations in dosing regimen of OM-85 administration (Fig. 3A-D) or in the choice of the endpoint (Fig. 3E-H) whereas the impact of SoC treatments parameters (duration of the TCS induction phase, TCS potency, and TCS administration frequency) appear to be negligible (Supplementary Information S2.1).

**Figure 3.**
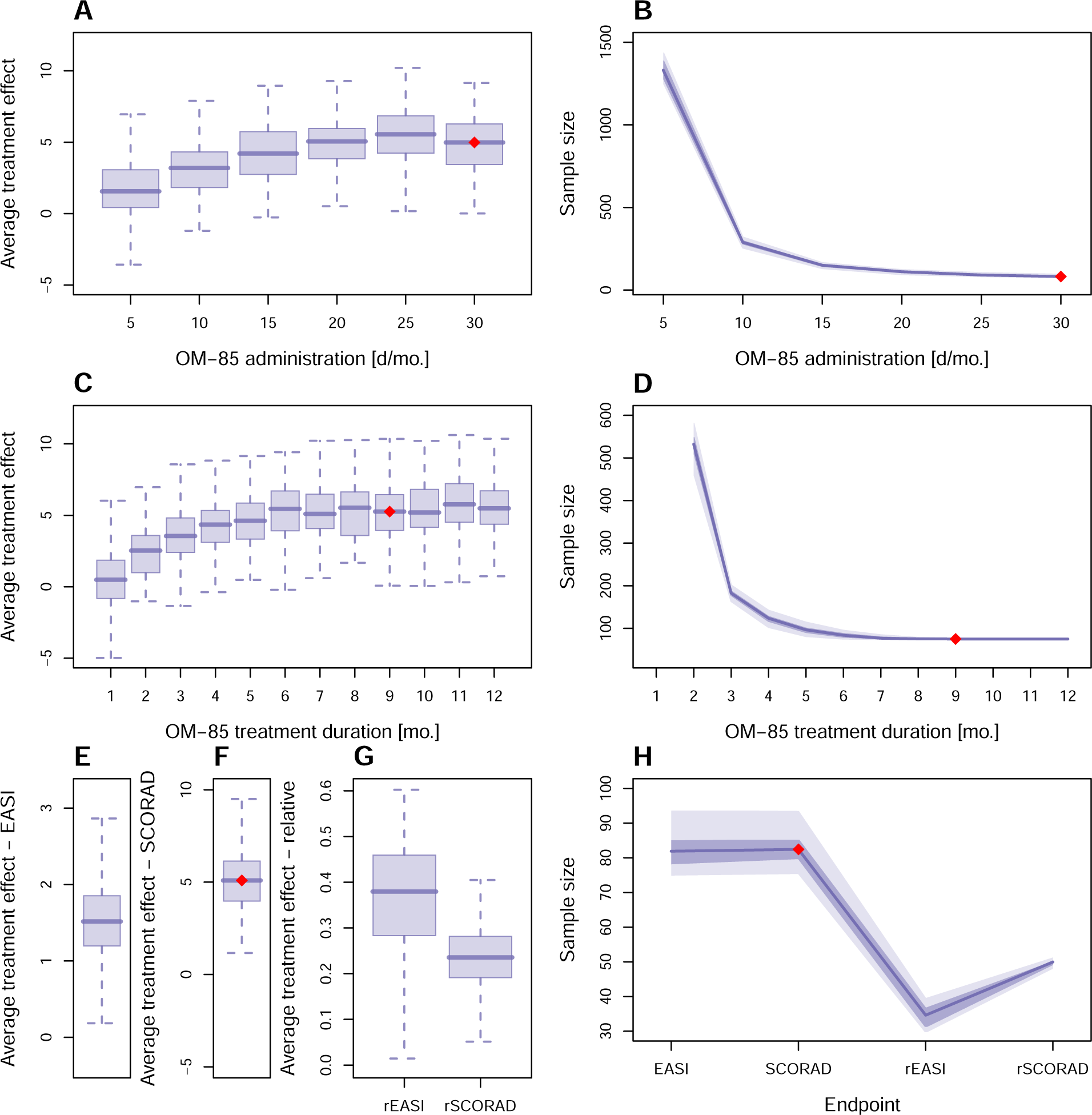
Sensitivity of trial results to trial protocol. Impact of (A – B) OM-85 administration frequency, from 5 to 30 days / month with a 5-day incremental step; (C – D) OM-85 treatment duration, from 1 to 12 months with a 1-month incremental step; (E – H) Endpoint: EASI, SCORAD, relative (*i.e.* normalized by the baseline severity) EASI (rEASI) and SCORAD (rSCORAD). The red dot represents the clinical trial settings of the study of Bodemer et al. ^28^.

The average treatment effect increases with the frequency of OM-85 administration with the best performance (Fig. 3 A: average treatment effect: 5, B. sample size: 83) being reached with a daily OM-85 administration as in Bodemer et al. ^28^. Compared with 5 days of OM-85 administration per month, the average treatment effect is 2 fold higher and the sample size 17 fold smaller. The efficacy also increases with the treatment duration until 6 months of treatment before reaching a plateau (Fig. 3 C: plateau average treatment effect 5.4, D: plateau average sample size 77). Compared with a single month of treatment, the average treatment effect is 10 fold higher and the sample size 6 fold smaller. Finally, our results indicate that the best endpoint is the relative EASI (Fig. 3E-G) for which the power (Fig. 3H) and sample size (Fig. 3I) are minimal: 34 patients are required compared with 83 when using absolute SCORAD. Note that average treatment effect appears to be more sensitive to the choice of an absolute vs relative endpoint than to the choice of scoring system (SCORAD or EASI, Fig. 3E).

### 3.3 QSP model-assisted early biomarker identification

In this section, we showcase how our QSP model can support a biomarker program early in clinical development with the aim of tailoring the target population by inclusion/exclusion criteria. This strategy is relatively common in AD: for example, focusing on higher disease severity strata can reduce population heterogeneity and potentially increase measured clinical benefit (explained e.g. by a larger need for treatment) but at the expense of targeting a more narrow subpopulation of AD patients^1^. Selection by phenotype and/or biomarkers may also help with treatment effect stratification and development of companion diagnostics already had success in other disease areas, e.g. in targeted cancer therapy^83^. However in the context of AD, the identification of predictive biomarkers seems to be difficult and to date, stratification by phenotypes (or associated markers) was not yet successful in stratifying treatment effect^84^ and only a handful of treatment-specific molecular biomarkers have been suggested so far with targeted therapies (see^85^. In part, this is due to the fact that identification of biomarkers predictive of the treatment effect requires the existence of rich datasets which include treatment effect and a panel of marker candidates. In the absence of such data, QSP models, acknowledged by regulatory bodies^86^ (especially in pediatric populations), have been suggested as promising tools to guide biomarker identification because they can leverage knowledge about the involved pathophysiological mechanisms thereby encoding information on how disease pathways respond to biomarker change.

#### 3.3.1 Enriched synthetic trial data generation

We make use of the following two capabilities of the disease model: a) the uncorrelated random variations in the 13 immune biomarker dimensions (Supplementary Information Table S4) in the virtual population which are in line with known marker variability and correspond to a broad diversity of AD phenotypes b) the alignment of the in silico clinical trial with historical trial data (Section 3.1). This allowed us to predict hypothetical, but biologically plausible, individual treatment effect for each virtual patient after OM-85 treatment including detailed immune cellular and soluble marker dynamics. This procedure augmented the existing demographics and disease severity data into a synthetic but highly immunologically enriched individual patient data set (Fig. 2). In the following sections, we then used this synthetic data to investigate various possible biomarker strategies for treatment effect stratification. Theoretically, there exists a range of options: from the use of disease severity strata to the use of thresholds on baseline markers levels (e.g. informed by a classifier based on linear or logistic regression and receiver operating curve validation) or of entire panels of quantitative marker levels (e.g. using treatment effect predicted from a model). This last strategy is known from other disease areas: for example, many metabolic disease trials employed stratification by insuline resistance as defined by HOMA-IR which uses fasting insulin and glucose levels and which was originally formulated as a rather complex non-linear ODE model^87^.

#### 3.3.2 Disease severity shows only limited potential for treatment effect stratification

In line with the common practice of selecting AD trial populations by severity, we investigated if baseline severity was a good predictor for the OM-85 effect in the reproduced and enriched reference trial data. As a first simple test, we performed a linear regression of the OM-85 effect with respect to virtual patient’s baseline disease severity, which resulted in a poor fit (R^2^ = 0.29). In order to exclude that the too simplistic picture of a linear regression confounds a severity stratification effect, we also employed our QSP model, but using a simplified virtual population where only individual baseline disease severity is varied (Supplementary Information S2.2). To better explain this approach, an analogy to mixed effect modeling^88^ is useful: in the reference virtual population, severity would be a random effect while all biomarkers would be mixed (with a severity-dependent mean and a random component). In the simplified severity-only population, all biomarkers’ fixed effects are kept but the random components are discarded. This approach performs poorly in comparison to the treatment effect values from our synthetic reference data (R^2^ = 0.35,Supplementary Information Figure S8.A), similarly to the linear regression. In line with that result, selecting a more severe patient subpopulation in our synthetic reference data is associated with slightly increased mean average treatment effect (Supplementary Information Figure S8.C1) as well as a predicted smaller sample size (Supplementary Information Figure S8.C2), but the restriction of the eligible patient fraction grows exponentially with the selection criteria (Supplementary Information Figure S8.C3). Selecting patients by baseline disease severity is therefore not the most promising avenue to responder enrichment in this example. This interpretation, however, has to be considered with caution since even though restricting the target population might be acceptable in some cases, clinical management aspects may become limiting for patient selection and more severe AD may have distinct pathophysiological mechanisms.

#### 3.3.3 Workflow for using our QSP model for low-dimensionality biomarker strategy

Consequently, in view of the unsatisfactory results of stratification by disease severity we therefore proceeded to take into account the coupled and non-linear nature of biomarker dynamics implemented into our QSP model. While in principle one could use the QSP model to predict treatment effect (and other output variables) using all biomarkers as inputs, this strategy may not be realistic in practice. Indeed, the need of comprehensive data to provide a value for all inputs is in contrast with the ethical burden of taking patient samples as well as economical considerations (e.g. extensive proteomic analyses^89^). Furthermore, the decision of committing to such a large data acquisition plan is difficult to take without some confidence in the fact that these data could efficiently stratify the patient population. Specific parts of our treatment-agnostic model may be less important for the effect of a specific treatment (such as OM-85, which acts via increase of the regulatory response, notably). Therefore, we sought a more parsimonous approach.

A preliminary investigation using typical model reduction technique fitting a series of generalized linear models (R^2^= 0.2, Supplementary Information S2.3 and Supplementary Information Figure S9.A) did not show promising trade-off between simplification potential and accuracy when simplifying the QSP model structure. We therefore aspired to develop a workflow where not the QSP model, but the overall in silico trial approach is simplified. For this, we reduced the virtual population by replacing some biomarker values by reference values (which depends on disease severity) with the idea of not drastically altering treatment effect, the primary outcome. Again, with a mixed effect modeling paradigm in mind, this amounts to removing as many random effects as possible without degrading the model’s goodness of fit. Such dimensional reduction approach has several advantages: a) it increases the chances for validation with experimental data and b) it enables the integration with a biomarker identification program in a realistic clinical setting with a limited number of biomarkers per test. This strategy is similar to feature selection techniques that have been suggested earlier to interface machine learning with QSP^90^.

Using the example of OM-85 versus placebo as add-on therapy on top of TCS and emollients, we therefore analyzed the contribution of the set of inputs in treatment effect variability. The result of this is the ranked list of markers that need to be supplied to the model (either from clinical data or the virtual population as a plausible theoretical counterpart of such data) in order to predict treatment effect. In practice, we performed an ANOVA-based global sensitivity analysis^91;92^ of treatment effect to baseline biomarkers using a virtual population of 2000 patients characterized by a unique baseline SCORAD (Supplementary Information Figure S11), which allowed to quantify the main effect of each biomarker and the effects of up to between four biomarker interactions. Ordering the biomarkers by their total effect (biomarker alone and sum of the interactions with any other biomarker) shows that each of the top four - alarmins, IL-22 cytokines, regulatory cytokines, and type-2 cytokines - explains more than 20% of the variance (and 83% in total) (Fig. 4B). Based on their main effect only (and not higher order interactions), they also exhibit the highest influence (8 to 15% each, 46% in total Supplementary Information Figure S12.A). We thus selected these lead biomarkers to predict treatment effect in the reduced input dimensionality approach. We then evaluated how this input dimensionality reduction approach performs, by confronting treatment effect values predicted with the reduced-variability vs the reference virtual population (Fig. 4A). While this approach leads to slight overestimation of treatment effect (Fig. 4C), the accuracy of the predictions is satisfactory (Fig. 4D, RMSE = 2.6, R^2^ = 0.63). Indeed, the prediction error (limited vs. full approach) is lower than 5 (SCORAD) for 94% of the virtual patients and the average error is 1.1 (min: 0.0002, Q1: 0.45, Q3: 2.23, max: 24.5). This prediction error is notably lower than measurement errors typically reported (normally distributed with a SD of 5^93^).

**Figure 4.**
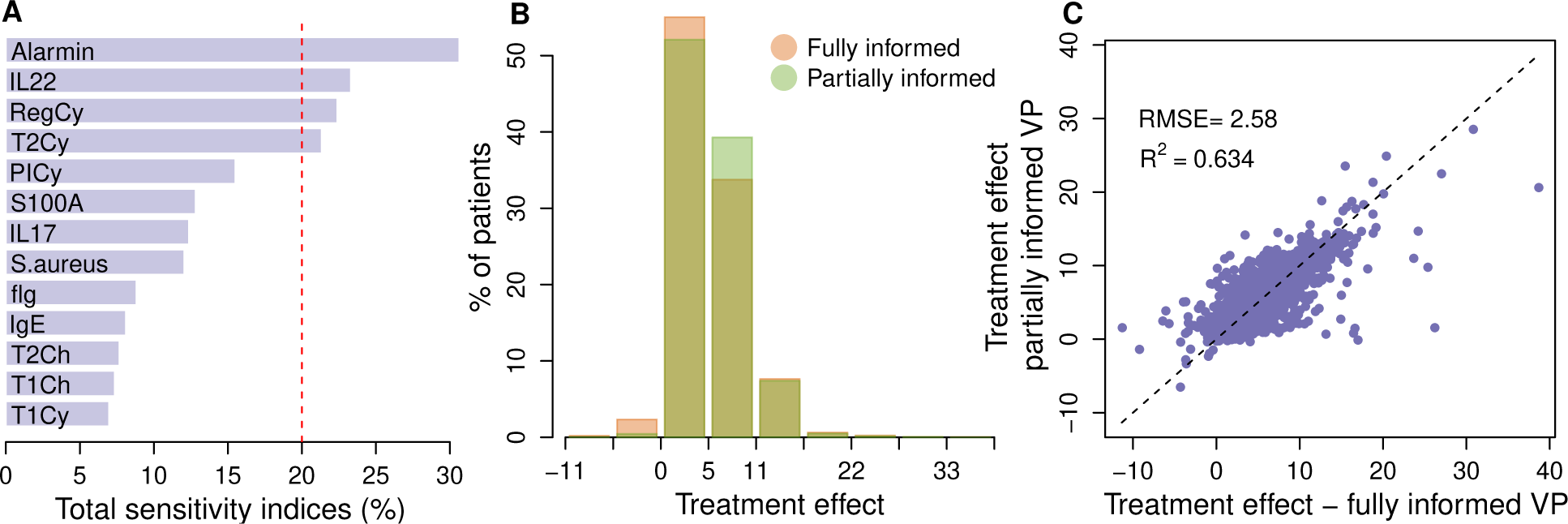
Low-dimensionality biomarker strategy. A. Global sensitivity analysis quantifying the (total: main + interactions) influence of baseline biomarkers on the treatment effect variance; selected key biomarkers have a sensitivity index above the red dashed line (20 %). B. Treatment effect distribution from the fully vs partially informed virtual population. C. Performance of the dimensionality reduction approach: comparison of treatment effect predicted by the fully vs partially informed virtual population.

We finally assessed the potential of using the biomarkers identified with the input dimensionality reduction approach for trial protocol optimization. In particular, we tested the effect of selecting virtual patients with required increasingly high treatment effect (predicted upon baseline marker selection); and we constructed subpopulations (including all patients – without selection – to the top 5% responders in incremental steps of 5%). For assessing this strategys performance, we report three indicators in Fig. 5: average treatment effect, sample size and statistical power. Selecting the top 50% responders results in 50% improvement of the average treatment effect (Fig. 5A), 50% reduction of the average sample size (Fig. 5B), and 31% improvement of the average chance of success of the trial (Fig. 5C). The maximal improvement for the average treatment effect (with respect to reference trial, enrolling only the top 5% predicted responders) is roughly 150% (Fig. 5A), the maximal reduction of the average sample size is 75% (Fig. 5B), and the maximal improvement of the average chance of success 33% (Fig. 5C). We also evaluated the impact of patient selection on the recruitment effort, defined as the total number of patients to be screened (Fig. 5D). The recruitment effort can be seen as a metric of the optimal selection criteria, as in the clinics, part of the cost would scale with the number of patients to screen. Our results suggest the optimal selection criterion lies between 25% and 60% of best responders, where the recruitment effort is the lowest, with a 19% average decrease compared to no selection. The recruitment effort exponentially increases with the selection stringency when selecting the top patients up to the top 60%, representing up to 300 patients to be screened for the top 5% selection criterion, *i.e.* 275% more than without selection.

**Figure 5.**
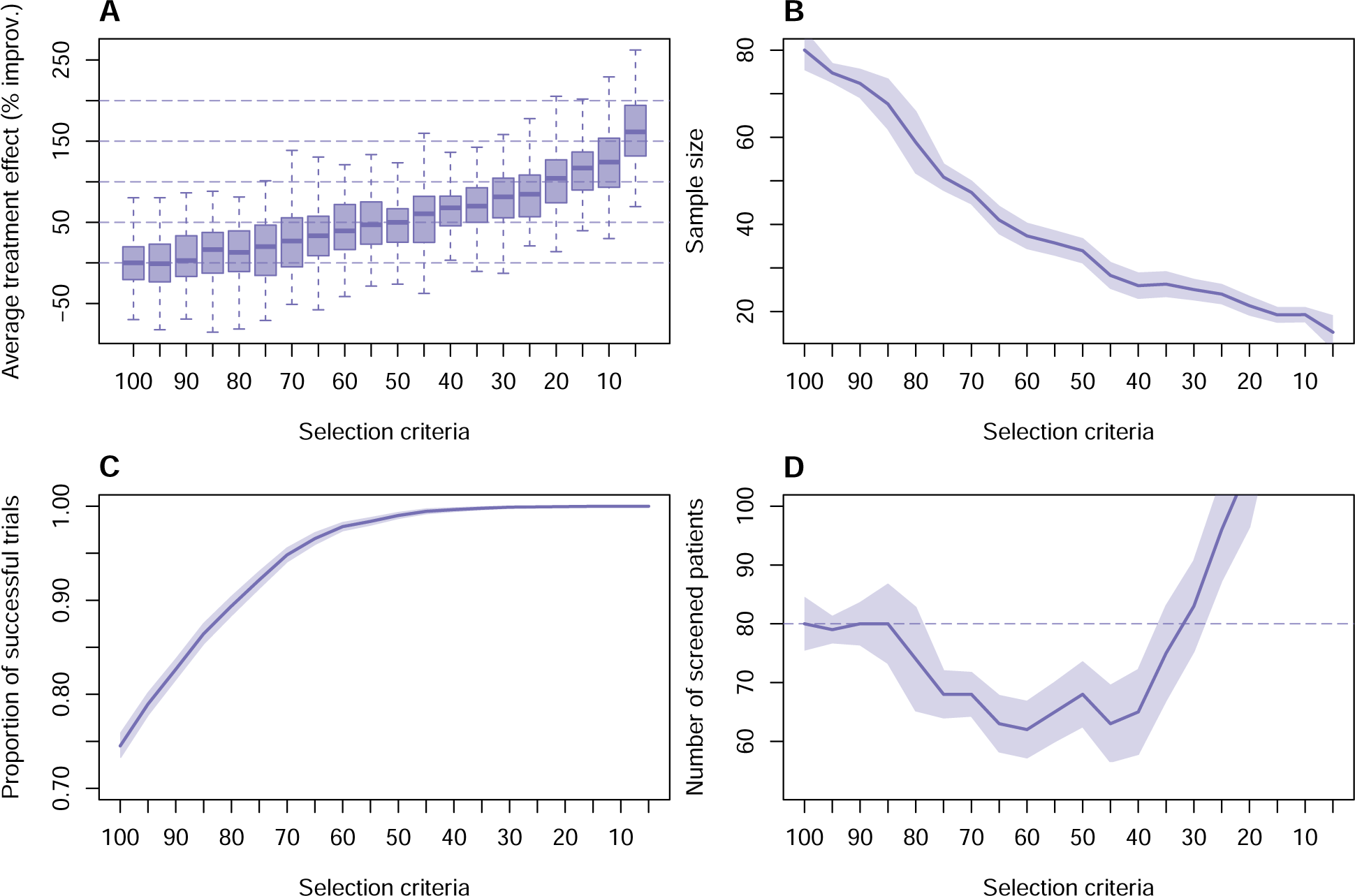
Clinical trial optimization based on predicted responders selection. A. Average treatment effect improvement (in %) vs increasingly strict selection criteria (top % responders): median (line), interquartile range (box), whisker (dashed line). B. Sample size distribution (obtained by bootstrapping over 100 samples) vs increasingly strict selection criteria: mean (line) and standard deviation (colored area). C. Statistical power (*i.e.* proportion of successful trials) distribution (obtained by bootstrapping over 100 samples) vs increasingly strict selection criteria: mean (line) and standard deviation (colored area). D. Recruitment effort (total number of patients to be screened during the recruitment process) distribution) vs increasingly strict selection criteria: mean (line) and standard deviation (colored area). Note that the y-axes have been reduced to highlight the optimum area, on average, 305 patients are to be screened for the selection of the top 5% best responders.

## 4 Discussion

### 4.1 Opportunities and challenges of combining complex QSP models and trial simulation

PBPK and PD models, QSP models, exposure response modeling and trial simulations are often regarded as distinct modeling paradigms under the umbrella of model-informed drug development^94^. There exist to date only few integrated modeling frameworks which couple QSP models and trial simulations^18;95–97^, most probably because they are complex to set up, multiscale by design, require a multi-stakeholder engagement and considerable effort for verification and validation^98;99^. One of the main hurdles for complex models (e.g. in QSP) is the high-dimensional parameter space which needs to be informed or calibrated. High complexity can be a necessary feature^100^ of the model and can also increase its robustness: different biological contexts can be covered and thus heterogeneous data from multiple sources can be used for calibration. On the other hand, high-dimensional models are prone to identifiability issues and parameter estimation suffers from non-convex and multi-modal objective functions with gradients that are computationally expensive to evaluate^101^. To add to this complexity, there is no consensus on the best method, and performance of such methods are model-specific^101^. We have therefore chosen an approach that minimized the number of parameters to calibrate. We have coupled our disease model, our SoC treatments models, and the treatment model of the immunomodulator OM-85^29^ into a QSP model. Central to to the disease model setup has been the choice of an equilibrium assumption (i.e. defining the system in terms of stationary dynamics), which allowed for calculating rather than calibrating a large fraction of the free parameters. In this way, from the set of 139 parameters present in the final QSP model, 70 could be constrained by the equilibrium approach; with 46 others being fixed with information from the literature, effectively leaving only 23 unknown parameters for calibration (i.e. with SoC data). The drawback of this method is that the pathogenesis, natural evolution and resolution of the disease are not described and that the used datasets (here skin proteomics in pediatric patients) convey context specificity (e.g. the model would need to be re-informed for adults and serum biomarkers).

While such a QSP model can resolve several questions around patient profiles and dosing-regimen, simulation studies of entire clinical trials needs a dedicated workflow. We therefore set up a virtual population of AD patients to reproduce the between-patient variability of our reference clinical trial^28^. We used arm designs (intervention, dosing-regimen), eligibility criteria and trial duration to perform trial simulations to predict drug efficacy according to a variety of trial protocols, therefore covering a wide spectrum of potential variables for trial design. Classically, in trial simulation used for statistical design, efficacy is considered as a constant by aggregating efficacy across all considered historical trials. However, when data exhibit heterogeneous treatment effects, uncertainty needs to be acknowledged and variability assessed by meta-regression and model-based meta-analysis. In fact, our QSP workflow can be considered as a corner case of model-based meta-analysis where several inputs from patient characteristics and posology are translated through mechanistic relationships into modulation of the efficacy. By using Monte-Carlo trial simulations (sometimes termed microsimulations) for several protocols and by assessing resulting simulation data for sample and effect size, we found a reduced sensitivity of efficacy optimization to dosing regimen (meaning that the dosing scheme from the reference trial is probably already optimal) compared to the choice of a relative severity endpoint over an absolute one which seems to harbor more potential for trial design optimization. In addition, this framework can be readily used for follow-up investigations like recruitment or cost-effectiveness considerations. One of the drawbacks of such trial simulation method (as compared to Monte Carlo simulation with a constant drug effect) is the high computational cost (per virtual patient) and the large storage and memory requirements when simulating many virtual patients.

### 4.2 Towards the holy grail of model-informed predictive biomarker exploration

Adjusting the dose and the treatment to each patient’s individual characteristics are the guiding principles of personalized medicine. Yet, today the gold standard for the approval of new drugs by regulatory agencies are placebo-controlled randomized clinical trials that report efficacy of entire populations of patients (see our discussion in Courcelles et al. ^102^ from a health technology assessment perspective). A way to better individualize drug development could be to use patient stratification and subgroup analysis, optimally performed on the basis of easily measurable biomarkers as objective (and ideally validated) predictors of treatment effect (predictive biomarkers). In fact, trials using biomarkers have an almost doubled overall probability of success compared to trials without biomarkers, notably in Phase I and II according to an analysis of Wong et al. ^103^, but the use of biomarkers outside of oncology is not very frequent^103^. To be implemented early-on, exploration of potential biomarkers has to be started prior to Phase I trials so that validation and qualification of biomarkers can be tackled during clinical development^104^. While in targeted oncology, markers (*i.e.* mutations) indicating the diseases vulnerability to treatment modulation are straightforward in their interpretation, inflammatory diseases often involve an intricate network of immune signaling and cellular pathways so that the search for biomarkers signatures is like a needle in the haystack^85^. In AD, progress has been made to identify biomarkers of severity such as systemic levels of the chemokine C-C motif ligand 17/thymus, an activation-regulated chemokine and chemoattractant of Th2 cells, which shows robust correlation with AD clinical severity, at both baseline and during therapy and thus could be used an objective surrogate for treatment effect^105^, but if such biomarker can also predict treatment effect for a specific investigational drug of interest is still an open question. The motivation behind this study was to use our QSP workflow to map the body of evidence of the relationships between biomarker levels and disease severity, in line with the immune dysregulation system involved in AD, onto a reference trial so that synthetic trial data is enriched with such biologically plausible information. As a proof of concept and to increase the practical applicability of our workflow, we tested several approaches to identify biomarkers predictive of treatment effect. We identified that neither a linearized surrogate model approach nor subgroup analysis by severity only lead to sufficient stratification potential, which a) plausibly underlines the difficulty to find predictive biomarkers in the context of AD (clinical data are in fact often analyzed using these statistical techniques), and b) emphasizes the need for methods which take into account non-linear effects for this purpose. Only an input dimensionality reduction approach, based on global sensitivity analysis proved accurate enough for identification of the skin biomarkers most predictive of OM-85 treatment effect. We did not intend to oversimplify the structural model (e.g. by lumping techniques) in view of the anticipated unbiased exploratory nature of the model. As linearization of the model was unsuccessful, the only viable option for simplification was to reduce the number of predictive input baseline biomarkers characterizing each virtual patient by replacing variability by fixed reference values. Quantification of how the input dimensionality reduction approach performs for predicting individual treatment effect makes us confident that embedding it within the *in silico* clinical trial framework can yield a treatment-specific tool that captures the state-of-the-art knowledge about how skin biomarkers are involved in the mechanism of action. As a perspective, this could a) inform the inclusion into the sample collection and analysis plan of e.g. Phase II trials, b) be prospectively validated through comparison with the generated clinical data and c) once qualified could support the clinical evidence by enrichment in sparsely sampled regions of biomarker space. For the use-case of OM-85, we found a subset of four skin biomarkers (alarmins, IL-22, type-2 and regulatory cytokines) predictive of treatment effect and we simulated that patient selection based on this subset may lead to larger effect size. In line with our results and among the skin biomarkers the model identified to be predictive of OM-85 effect, IL-22 and IL-13 expression levels in the skin tissues have been previously identified to strongly and significantly correlate with clinical therapeutic effect in AD^105^. Development of such computational approaches goes well in hand with promoting clinical trials designed to validate or reject the predictions made by the model and which could then either be used to improve subsequent (e.g. Phase IIb and III) clinical trial designs or in a learn and confirm paradigm to refine our model^105^.

## Supporting information

Supplementary Methods, Results, Figures and Tables

## Data Availability

All information needed to reproduce the simulations are described in the references of this manuscript, and the supplementary information. Access to the computational model code, documentation and simulation results are available on Jinko.ai platform upon request to the corresponding author. R scripts developed for analysis of simulation results and producing the figures are also available upon resonable request.

## Abbreviations

AD: Atopic dermatitis
SoC: Standard of care
TCS: Topical corticosteroids
ODE: Ordinary differential equation
QSP: Quantitative systems pharmacology
PBPK: Physiologically based pharmacokinetic
PD: Pharmacodynamics
AB: Absolute benefit
SCORAD: Scoring atopic dermatitis
EASI: Eczema area and severity index

## Acknowledgments

We acknowledge Valentin Vierge and Daniel Šmít for contributing to model building and calibration, Germàn Gómez for statistical support, and Louis Philippe for technical support.

## Authors contributions

A.K., S.A., L.L., C.P and C.S. supervised the study. N.G., S.A. and A.K. designed the model. N.G, S.A, I.F., T.G., S.M.B., D.L., Y.W., L.E. developed the model. N.G. performed the simulations and analyzed the results. N.G., S.A. and A.K wrote the manuscript. All authors contributed to the discussion of the results and reviewed the manuscript.

## Supplementary Information

The Supplementary File contains: (S1) Supplementary Methods where the (S1.1) Model, (S.1.2) Calibration, (S1.3 Biomarker selection, and (S1.4) Bootstrapping procedure are described in details; (S2) Supplementary Results for (S2.1) Assessment of the efficacy sensitivity to the TCS related trial protocol settings, (S2.2) Performance baseline AD severity as a marker of treatment effect, and (S2.3) Identification of predictive biomarkers using a surrogate model approach; (S3) Supplementary Figures and (S4) Supplementary Tables.

