## Supplementary Methods, Results, Figures and Tables for "A quantitative systems pharmacology workflow towards optimal design and biomarker stratification of atopic dermatitis clinical trials"

#### Abstract:

*Background.* The development of atopic dermatitis (AD) drugs is confronted by many disease phenotypes and trial design options, which are hard to explore experimentally.
*Objective.* Optimize AD trial design using simulations.
*Methods.* We constructed a quantitative systems pharmacology (QSP) model of AD and standard of care (SoC) treatments and generated a phenotypically diverse virtual population whose parameter distribution is a) derived from known relationships between AD biomarkers and disease severity and b) calibrated using disease severity evolution under SoC regimens.
*Results.* We applied this workflow to the immunomodulator OM-85, currently being investigated for its potential use in AD, and calibrated investigational treatment model with the efficacy profile of an existing trial (thereby enriching it with plausible marker levels and dynamics). We assessed the sensitivity of trial outcomes to trial protocol and found that for this particular example, a) the choice of endpoint is more important than the choice of dosing-regimen and b) patient selection by model-based responder enrichment could increase the expected effect size. A global sensitivity analysis reveals that only a limited subset of baseline biomarkers is needed to predict the drug response of the full virtual population
*Conclusion.* This AD QSP workflow built around knowledge of marker-severity relationships as well as SoC efficacy can be tailored to specific development cases so as to optimize several trial protocol parameters and biomarker-stratificaiton and therefore holds promise to become a powerful model-informed drug development tool.

**S3 Supplementary Figures** **22**

**S4 Supplementary Tables** **35**

**References** **43**

### S1 Supplementary methods

#### S1.1 Quantitative systems pharmacology model

We consider a representative sample of lesional atopic dermatitis (AD) skin of 5 cm$^3$. This corresponds to the volume of skin tissue defined by the surface typically sampled by tape stripping (about 2 cm$^2$) and an average skin thickness of 2.5cm. Generally, we disregard geometrical considerations regarding extension/receding of the lesion and consider only the severity of the lesion. We also consider the volume of lesional skin as a well-mixed compartment and disregard notion of depth and layers, which is a strong hypothesis given the highly structural nature of the skin as a tissue. In this section, we present the equations governing the dynamics of the biological entities constituting the model (Tab. S1). We describe the forces acting on their variations (positive or negative) shaped by the regulation relationships between them (Box S1). Notations used for the model parameters are presented in Tab. S2.

The model focuses primarily on the interplay between the skin barrier integrity and the skin immune system. Indeed, epidermal barrier dysfunction is consistently observed in affected and unaffected skin of patients with AD[1,2]. Disruption of skin barrier function in AD is reported to be a multifactorial process, which includes genetic factors, such as filaggrin (FLG) mutations, and physical damage from scratching[1]. AD affected skin is permissive to allergens and pathogens, including *Staphylococcus aureus* present in the skin microbiome (*S. aureus*)[3–5]. In return, colonization or infection with *S. aureus* damages the skin barrier[1,3,5], which induces type-2-skewed immune dysregulation and cutaneous inflammatory response[1,4,5]. In addition, the loss of skin barrier integrity also induces directly (release of alarmins) and indirectly (allowing pathogens, allergens and irritants to permeate) this type-2-skewed immune dysregulation[1]. In return, the type-2 immune activity in the skin exacerbates the underlying barrier defect[1,2,6].

##### S1.1.1 Skin barrier integrity

Skin barrier integrity ($S$) dynamics is modeled following a regeneration – degradation formalism, and the variable is normalized between 0: completely damaged skin and 1: healthy skin.

- *Regeneration* We set the skin barrier regeneration directly proportional (rate $r_S$) to the concentration of filaggrin protein ($F$). Indeed, it is well established that filaggrin proteins play a central role in the integrity of the skin barrier, and this via multiple mechanisms: ensuring the water holding capacity, maintenance of acidic pH, maintenance of the structural and mechanical integrity[10–16]. We additionally account for an amplification of the skin barrier regeneration in presence of IL-22 or regulatory cytokines ($\mathcal{R}_{+}^{S,Y}$). Indeed, several evidences suggest that IL-22 can promote epithelial cell proliferation, survival and repair in the skin as well as prevent tissue destruction[17]; and in vivo exploration of cytokine expression levels suggest that TGF-$\beta$ is involved in skin tissue remodelling[18,19]. Note that the regeneration term is multiplied by $(1-S)$ in order to constrain the variable between 0 and 1. Finally, we account for an additional term of skin regeneration (regulation $\mathcal{R}_{+}^{E,S}$) in presence of emollients[20–25].

- *Degradation* Skin barrier degradation is set to be driven by several cytokines ($\mathcal{R}_{-}^{S,Y}$). Indeed, multiple evidences suggest that pro-inflammatory[26], type-2 (IL-4 and IL-13)[1,3,6,27,28], and S100A[29] cytokines directly and indirectly exacerbate skin barrier dysfunctions, for instance by altering protein and lipid content. Note that IL-22 also has a negative effect on the skin barrier integrity[30], which we assume negligible compared with its positive effect. We also neglect the negative impact of type-1 cytokines[18,26] to ensure model stability, in particular to allow the model dynamics to return to the equilibrium values after treatment termination. Finally, *S. aureus* is known to (directly and indirectly) damage the skin barrier integrity[1,3,5], which is accounted for by an additional down-regulation function ($\mathcal{R}_{-}^{S,P_S}$).

Dynamics of skin barrier integrity ($S$) read:

**Box S1.** Regulations formalism

In immunological models, cytokine effects are classically represented based on a saturation formalism, e.g. the Hill function (see for example Wigginton et al. 2001[7], Gammack et al. 2005[8], Marino et al. 2010[9]), to reflect the fact there is a limited number of the modulator receptors on the cell surface, so the effect saturates above a given modulator concentration. Similarly, we based the formalism of our regulation on Hill functions: $\mathcal{R}^X$ formalize the regulation of a immune mechanisms $\mathcal{M}$ depending on the levels of the modulator $X$; in which $k_M^X$ denotes the (positive) half-saturation constant and $v_{max}^X$ denotes the (positive) saturation constant or maximum reaction rate. Note that regulations due to Standard of Care (SoC) do not follow this formalism (see dedicated part at the end on this box)

**Up-regulations** $\mathcal{R}_+^X$ is written as follows:

$$\mathcal{R}_+^X = \frac{v_{max}^X \times X^n}{\left(k_M^X\right)^n + X^n} \quad \text{where} \quad v_{max} > 0 \quad \text{and} \quad k_M > 0$$

In most cases the Hill coefficient are set to $n = 1$, the Hill function corresponds thus to a Michaelis-Menten formalism. With this formalism,

- at low concentration of the modulator $X$, we have $\mathcal{R}_+^{X \to 0} \to 0$
- at $X = k_M^X$, we have $\mathcal{R}_+^{X = k_M^X} = 0.5 \times v_{max}^X$
- at high concentration of the modulator $X$, we have $\mathcal{R}_+^{X \to \max(X)} \to v_{max}^X$

Up-regulations can be either activation or amplification of immune mechanism. Considering a given up-regulated mechanism $\mathcal{M}$, the activation *vs* amplification of the corresponding mechanism writes as follows:

- *activation*: $\mathcal{M} \times \mathcal{R}_+^X$, which means that with a zero concentration of the modulator $X$, the mechanism $\mathcal{M}$ is turned-off.
- *amplification*: $\mathcal{M} \times \left(1 + \mathcal{R}_+^X\right)$, which means that with a zero concentration of the modulator $X$, the mechanism $\mathcal{M}$ occurs at its basal rate.

In case of **multiple modulators** ($X_1$ and $X_2$) involved in the same regulation $\mathcal{R}_+^X$ of the mechanism $\mathcal{M}$, their effects are summed. In some cases, multiple modulators have a **synergistic effect**, represented by an interactive term:

$$\mathcal{M} \times \left(1 + \mathcal{R}_+^X\right) \quad \text{With} \quad \mathcal{R}_+^X = \frac{v_{max}^{X1} \times X_1}{k_M^{X1} + X_1} + \frac{v_{max}^{X2} \times X_2}{k_M^{X2} + X_2} + \frac{v_{max}^{X1,X2} \times X_1 \times X_2}{\left(k_M^{X1} + X_1\right) \times \left(k_M^{X2} + X_2\right)}$$

**Down-regulations** $\mathcal{R}_-^X$ also follow a Hill function formalism, where the saturation constant is assumed to be equal to one: $v_{max}^X = 1$ and is written as follows:

$$\mathcal{M} \times \left(1 - \frac{v_{max}^X \times X}{k_M^X + X}\right) = \mathcal{M} \times \frac{k_M^X}{k_M^X + X} = \mathcal{M} \times \mathcal{R}_-^X$$

With this formalism,

- at low concentration of the modulator $X$, we have $\mathcal{R}_-^{X \to 0} \to 1$
- at $X = k_M^X$, we have $\mathcal{R}_-^{X = k_M^X} = 0.5$
- at high concentration of the modulator $X$, we have $\mathcal{R}_-^{X \to \max(X)} \to 0$

In case of **multiple modulators** ($X_1$ and $X_2$) involved in the same down-regulation $\mathcal{R}_-^X$ of the mechanism $\mathcal{M}$, we defined the down-regulation as the product of the down-regulations by $X_1$ and by $X_2$ respectively:

$$\mathcal{M} \times \mathcal{R}_-^X = \mathcal{M} \times \mathcal{R}_-^{X1} \times \mathcal{R}_-^{X2} = \mathcal{M} \times \frac{k_M^{X1}}{k_M^{X1} + X_1} \times \frac{k_M^{X2}}{k_M^{X2} + X_2}$$

**Combining up- and down-regulations**

When up- and down-modulators of a same mechanism are present, then their effects are summed, with a few exceptions (notably for the regulations induced by TCS, see dedicated part below). For instance assuming the mechanism $\mathcal{M}$ amplified by $X_1$ and inhibited by $X_2$, we write $\mathcal{M} \times (1 + \mathcal{R}_+^{X1} + \mathcal{R}_-^{X2})$. As a consequence, we assume there is no competitive inhibition, *i.e.* the up- and down-modulators bind to different receptors.

**Exception: regulations due to SoC** Their effects on a given mechanism $\mathcal{M}$ is formalized via a term $\mathcal{R}^{SoC,\mathcal{M}}$ defined as the product of a constant (calibrated) $v_{max}^{SoC,\mathcal{M}}$, a binomial factor that turns-on / off the regulation in presence / absence of the SoC $\delta_{SoC}$, and for TCS only, a scaling factor linked to the potency of the TCS $s_p$. This is justified by the fact that we neglect the pharmacokinetic of SoC (see sec. S1.1.15).

$$\frac{\mathrm{d}S}{\mathrm{d}t} = r_S\,F\,\left(1 + \mathcal{R}_{+}^{S,Y}\right)\,(1 - S) + \mathcal{R}_{+}^{E,S} - \left(\mathcal{R}_{-}^{S,Y} + \mathcal{R}_{-}^{S,P_S}\right)\,S \tag{S1}$$

###### S1.1.2 Filaggrin levels

Filaggrin protein ($F$) dynamics is modeled by a production (rate $r_F$) – decay (rate $\mu_F$) formalism. Based on literature, filaggrin production is set to be inhibited by type-2[6,27] as well as type-17 and type-22[18,27] cytokines, via the down-regulation $\mathcal{R}_{-}^{F}$.

Dynamics of filaggrin protein levels reads:

$$\frac{\mathrm{d}F}{\mathrm{d}t} = r_F\,\left(1 + \mathcal{R}_{-}^{F}\right) - \mu_F\,F \tag{S2}$$

###### S1.1.3 *S. aureus*

As AD patients have a higher proportion of *S. aureus* in their skin microbiome compared with healthy controls[1,3,4], we approximate the skin microbiome by a variable representing the relative abundance of *S. aureus* in percentage ($P_S$). In order to constrain the variable between 0 and 100%, we based its dynamics on a carrying capacity formalism (intrinsic growth rate: $r_{P_S}$, carrying capacity $c_{P_S}$, decay rate: $\mu_{P_S}$). As *S. aureus* colonization was shown to be directly related to the severity of AD[3], we assume in our model its ability to colonize the microbiome to be limited by the skin barrier integrity (renewal term multiplied by $(1 - S)$). Furthermore, proteins with anti-microbial or host-defense functions have been suggested as part of the main drivers preventing *S. aureus* colonization[27,31]. We account for this factor via two regulations:

- Down-regulation $\mathcal{R}_{-}^{P_I}$ which represent the limitation of *S. aureus* colonization via immune proteins, namely S100A for its anti-microbial functions[32] and type-1 cytokines as drivers of the synthesis of $\beta$-defensin[33].
- Up-regulation $\mathcal{R}_{+}^{P_I}$, which represent a predisposing factor to *S. aureus* colonization via type-2 cytokines due to their ability to limit $\beta$-defensin expression[33].

Dynamics of *S. aureus* abundance in the skin microbiome read:

$$\frac{\mathrm{d}P_S}{\mathrm{d}t} = r_{P_S}\,P_S\,\left(1 - \frac{P_S}{c_{P_S}}\right)\,(1 - S)\,\left(1 + \mathcal{R}_{+}^{P_I} + \mathcal{R}_{-}^{P_I}\right) - \mu_{P_S}\,P_S \tag{S3}$$

###### S1.1.4 Pathogen infiltration

In our model, pathogens consist of a group of very different biological entities (virus, bacteria including *S. aureus*, allergens, etc) such that it can not be quantitatively informed. We define pathogen infiltration as a semi-quantitative variable ($P_I$) varying between 0 (healthy patient) and 1.

We model the dynamics of pathogen infiltration as (i) a contribution from the proportion of *S. aureus* (term $s_{P_S}^{P_I}\,P_S$) and (ii) an infiltration (rate $r_{P_I}$) – decay (rate $\mu_{P_I}$) formalism. With regards to the infiltration rate $r_{P_I}$, we define it as a product of the exposure to pathogens from the environment (which has been further constrained by the equilibrium conditions) and the skin permeability (arbitrarily set at the same value than the previously published model from Domínguez-H¨˜ttinger *et al.* (2017)[34]).

Furthermore,the pathogen infiltration is determined by the integrity level of the skin barrier (multiplied by $(1-S)$[35–37]. Finally, we apply the same up- ($\mathcal{R}_{+}^{P_I}$) and down- ($\mathcal{R}_{-}^{P_I}$) regulations of pathogen infiltration as for the dynamics of *S. aureus* colonization ((S3)). Additionally, we account for the neutralization of pathogens by polyclonal IgA ($A_A$)[38,39] under OM-85 treatment, formalized as an up-regulation of the infiltrated pathogen decay (regulation $\mathcal{R}_{+}^{\mu P_I, A_A}$).

As a result, dynamics of infiltrated pathogens read:

$$\frac{\mathrm{d}P_I}{\mathrm{d}t} = s_{P_S}^{P_I}\, P_S \,+\, r_{P_I}\,(1\,-\,S)\,\left(1\,+\,\mathcal{R}_{+}^{P_I}\,+\,\mathcal{R}_{-}^{P_I}\right)\,-\,\mu_{P_I}\,(1+\mathcal{R}_{+}^{\mu P_I, A_A})\,P_I \tag{S4}$$

##### S1.1.5 Keratinocytes

We set their number ($K$) in the given lesional AD skin sample to follow a positive linear relationship (rate $\frac{c_K}{2}$) with the skin barrier integrity ($S$) as keratinocytes are the major cell type of the epidermis[40]. In our model, we constrain their number to be between:

- in healthy skin: $K = c_K$ when $S = 1$, which was arbitrary set as no data was found.
- half of the number in healthy skin when skin is completely damaged: $K = 0.5 \times c_K$ when $S = 0$. Note that this remains an assumption as we found no data to inform the minimal number of keratinocytes.

Number of keratinocytes ($K$) reads:

$$K = \frac{c_K}{2}\,(1\,+\,S) \tag{S5}$$

##### S1.1.6 Type-2 Innate Lymphocyte Cells (ILC2)

Among Innate Lymphocyte Cells (ILCs), only ILC2 are considered to be important in the pathogenesis of AD[41]. This is why we focus on activated ILC2 ($L$). Their dynamics follows an activation (rate $\alpha_L$) – decay (rate $\mu_L$) formalism. Their activation is dependent on alarmins[41,42] (regulation $\mathcal{R}_{+}^{Y_A}$).

Dyanmics of ILC2 cells read:

$$\frac{\mathrm{d}L}{\mathrm{d}t} = \alpha_L\,\mathcal{R}_{+}^{Y_A}\,-\,\mu_L\,L \tag{S6}$$

##### S1.1.7 Dendritic cells

Dendritic cells (DC) population in atopic skin mainly consists of Langerhans cells (LC) and Inflammatory Dendritic Epidermal Cells (IDEC)[43]. We only explicitly modeled the activated and mature state of each DC population. This model simplification is based on exploratory analyses which showed that including the naive sate of both DC was associated with a strong predominance of activated over naive DC, and overall model predictions were comparable with and without representing naive DC dynamics. Dynamics of both LC ($D_L$) and IDEC ($D_E$) are modeled by an activation / recruitment – decay (rates $\mu_{D_L}$, $\mu_{D_E}$) formalism. Assuming a constant concentration of naive LC equal to $c_{D_L}$, LC are activated (at maximal rate $\alpha_{D_L}$) in response to alarmin signals[44] (regulation $\mathcal{R}_{+}^{Y_A}$). As IDEC are recruited in the skin tissues only in presence of pro-inflammatory cytokines[43,45,46], we set their activation (rate $r_{D_E}$) directly depending on the levels of pro-inflammatory cytokines ($Y_I$) via the regulation $\mathcal{R}_{+}^{D_E}$. We finally account for an up-regulation of DC death (regulation $\mathcal{R}_{+}^{C,\mu D}$) in presence of TCS[47–52].

Dynamics of LC ($D_L$) and IDEC ($D_E$) read:

$$\begin{aligned}\frac{\mathrm{d}D_L}{\mathrm{d}t} =&\alpha_{D_L}\, c_{D_L}\, \left(\mathcal{R}_{+}^{Y_A}\right) - \mu_{D_L}\, (1 + \mathcal{R}_{+}^{C,\mu D})\, D_L \\ \frac{\mathrm{d}D_E}{\mathrm{d}t} =&r_{D_E}\, \mathcal{R}_{+}^{D_E} - \mu_{D_E}\, (1 + \mathcal{R}_{+}^{C,\mu D})\, D_E\end{aligned} \tag{S7}$$

##### S1.1.8 Type-1 innate memory like cells

Mirroring the mechanisms of action of OM-85 in the context of respiratory tract infections[53], we assume the homing in the skin tissues of type-1 innate memory like cells $D_M$ from the systemic circulation under OM-85 treatment. Generation and trafficking of these cells is set as in the already published model from Arsène *et al.* (2022)[53]. The only difference is their infiltration in skin tissues as well, but which is implemented in our model following the same formalism:

$$\begin{aligned}\frac{\mathrm{d}D_M}{\mathrm{d}t} =&\varphi\, (1-\sigma)\, D_M^v - \mu_{D_M}\, D_M \\ &\varphi \quad \text{the lymphatic flow in the skin} \\ &\sigma \quad \text{the vascular reflection coefficient} \\ &D_M^v \quad \text{the concentration of type-1 innate memory like cells in the systemic circulation}\end{aligned} \tag{S8}$$

##### S1.1.9 Regulatory T-cells (T-reg)

Not a lot is known on the mechanisms driving the dynamics of regulatory T-cells in lesional AD skin[6,54]. Depending on their phenotype, they would either express pro-inflammatory or regulatory functions[6]. Furthermore, T-cell types could exhibit phenotype plasticity depending on the cytokine environment, for instance conversion of T-reg cells into Th2 cells would occur[54]. Given the paucity of knowledge, we decided to focus in our model on T-reg exhibiting regulatory functions and we neglect T-cell plasticity. We thus set T-reg dynamics as following a minimalist production (rate $r_{T_R}$) – decay (rate $\mu_{T_R}$) formalism.

Mirroring the mechanisms of action of OM-85 in the context of respiratory tract infections[53], we assume the homing in the skin tissues of regulatory T-cells from the systemic circulation under OM-85 treatment. Generation and trafficking of these cells is set as in the already published model from Arsène *et al.* (2022)[53]. The only difference is their infiltration in skin tissues as well, but which is implemented in our model following the same formalism (term $r_{T_R}^v$, see also Equation S8)

Dynamics of regulatory T-cells read:

$$\frac{\mathrm{d}T_R}{\mathrm{d}t} =r_{T_R} + r_{T_R}^v - \mu_{T_R}\, T_R \qquad \text{Where } r_{T_R}^v = \varphi\, (1-\sigma)\, T_R^v \tag{S9}$$

##### S1.1.10 Chemokines

We model the dynamics of the major type-1 ($H_1$) and type-2 ($H_2$) chemokines in the context of AD. In our model, type-1 chemokines consist of CXCL10, which has been shown to be up-regulated in AD skin of adult patients at least[55–58]. In our model, type-2 chemokines group CCL17, CCL22, and CCL27. We decided to account for CCL7 and CCL22 because they are part of the main biomarkers of the AD systemic response and strongly and significantly correlate with patient response[55]; and for CCL27 (CTACK) because it is expressed only in the skin[59] and allows the distinction between AD and psoriasis[55]. The dynamics of type-1 chemokines $H_1$, and type-2 chemokines $H_2$ are set to follow a production (rates $\rho_{H_1}$ and $\rho_{H_2}$) – decay (rates $\mu_{H_1}$ and $\mu_{H_2}$) formalism (Equations (S10) and (S11) respectively).

The production term of both chemokine types is set as driven by LC and IDEC as they are both known to release type-1[60] and type-2 chemokines[61]. We additionally implemented three regulations for this production term, which is set to be:

- amplified when antigen uptake via the Fc$\varepsilon$RI – IgE complex on the DC surface occur (regulation $\mathcal{R}_{+}^{\rho D}$), as suggested by the literature knowledge[43,45]. For this we define $\mathcal{R}_{prod}^{\rho D} := D_L \left(1+\mathcal{R}_{+}^{\rho D}\right) + D_E \left(1+s_{D_L}^{D_E}\,\mathcal{R}_{+}^{\rho D}\right)$ (Box S2).
- dependent of the cytokine environment, either pro type-1 for the release of type-1 chemokines (regulation $\mathcal{R}_{env}^{H_1}$) or pro type-2 for the release or type 2 chemokines (regulation $\mathcal{R}_{env}^{H_2}$). This relies on the generalization of the finding that CCL17 expression is induced by the type-2 cytokines while down-regulated by type-1 cytokines[61].
- inhibited by regulatory cytokines (regulation $\mathcal{R}_{-}^{Y_R}$), based on the literature knowledge suggesting that regulatory cytokines limit both type-1 and type-2 responses[2,18,54].

Dynamics of type-1 chemokines ($H_1$) read:

$$\frac{\mathrm{d}H_1}{\mathrm{d}t} = \rho_{H_1}\,\mathcal{R}_{prod}^{\rho D}\,\mathcal{R}_{env}^{H_1}\,(1\;+\;\mathcal{R}_{-}^{Y_R}) - \mu_{H_1}\,H_1 \tag{S10}$$

In addition, type-2 chemokines are also released by keratinocytes, and this production is amplified by pro-inflammatory and type-2 cytokines[59,61,62]. Hence, we set an additional production term of type-2 chemokines as follows: $\mathcal{R}_{prod}^{K} = K\,(1-S)\,\left(1+\mathcal{R}_{+}^{K}\right)$

Dynamics of type-2 chemokines ($H_2$) read:

$$\frac{\mathrm{d}H_2}{\mathrm{d}t} = \rho_{H_2}\,\left(\mathcal{R}_{prod}^{K} + \mathcal{R}_{prod}^{\rho D}\,\mathcal{R}_{env}^{H_2}\,(1\;+\;\mathcal{R}_{-}^{Y_R})\right) - \mu_{H_2}\,H_2 \tag{S11}$$

##### S1.1.11 Cytokines

In our model, we represent the dynamics of the main cytokines involved in AD but rather than representing the dynamics of each cytokines, we decided to group them by functional groups, as follows:

ALARMINS ($Y_A$) grouping thymic stromal lymphopoietin (TSLP), IL-25, and IL-33[3,6,42,45]

PRO-INFLAMMATORY ($Y_I$) grouping IL-1$\beta$, IL-6, IL-8, and TNF-$\alpha$[60,65]

**Box S2.** Modeling formalism of DC immune functions induced by antigen uptake

*Biological mechanisms* Pathogens carry antigens that can bind to dendritic cells via IgE-FceRI complexes[43], which further induces (or amplified) the expression of DC immune functions: chemokines synthesis[43,45] and antigen presentation to the adaptive immune response resulting in the release of adaptive-derived cytokines[43,63] (associated Figure). Antigens uptake by the IgE – Fc$\varepsilon$RI binding increases the surface expression of this complex[45] (Figure below). Furthermore, its expression of Fc$\varepsilon$RI is amplified by alarmin signals[64] (Figure below).

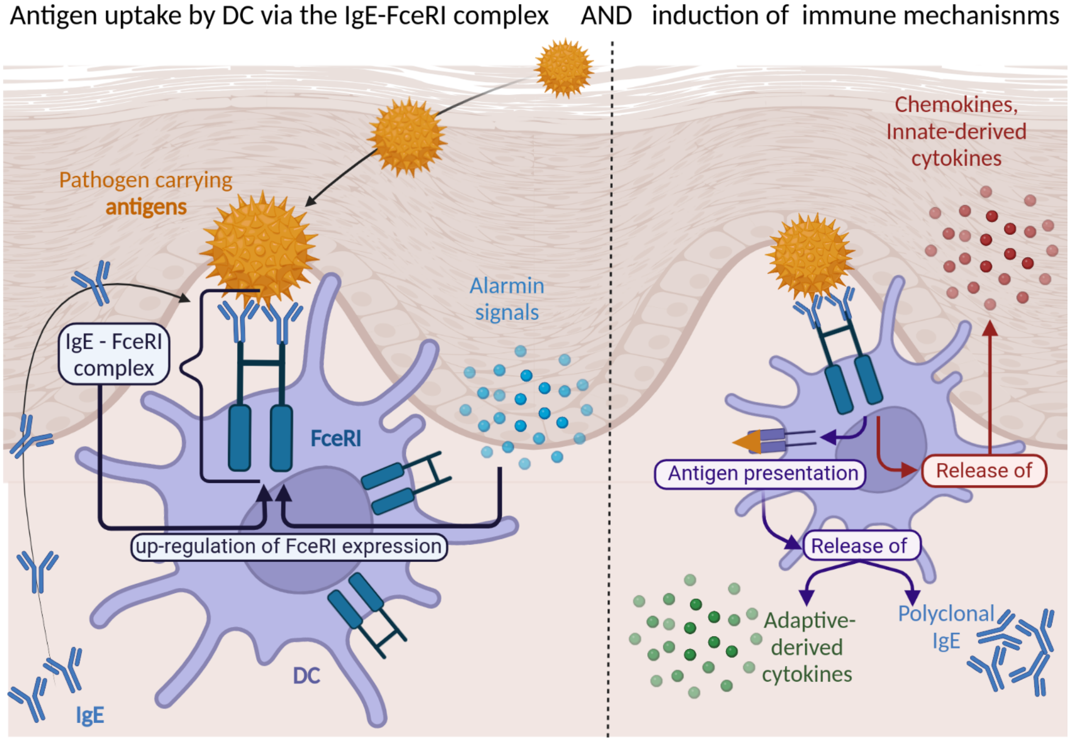

*Formalism* We do not explicit the dynamics of Fc$\varepsilon$RI expression on DC surface but we account for immune functions induced by antigen uptake via IgE – Fc$\varepsilon$RI complexes via the regulation $\mathcal{R}_+^{\rho D}$. Here, we demonstrate that this simplification does not relies on strong assumptions.

First, lets define $\mathcal{R}_+^{\rho D}$ the up-regulation of DC immune functions due to antigen uptake, which can we written as follows (see Box S1): $\mathcal{R}_+^{\rho D} = \frac{v_{max}^{P_I}(P_I)^n}{(k_M^{\rho D})^n + (P_I)^n}$; where $P_I$ is the concentration of pathogens in the skin tissues, and $n$ is set to 2. $v_{max}^{P_I}$ represents the saturation constant and can be linked to to number of Fc$\varepsilon$RI complexes expressed at the surface of DC ($D$). Thus, we can define $v_{max}^{P_I}$ as a function of Fc$\varepsilon$RI: $v_{max}^{P_I} = f(\varepsilon)$. To inform this function, we can write the dynamics of Fc$\varepsilon$RI in the lesional skin, based on an expression (rate $\rho_\varepsilon$) – degradation (rate $\mu_\varepsilon$) formalism, accounting for the amplification of its expression depending of IgE antibodies ($A_E$) and alarmin signals ($Y_A$): $\frac{d\varepsilon}{dt} = \rho_\varepsilon\, D\left(\frac{Y_A}{k_M^{Y_A}+Y_A} + \frac{A_E}{k_M^{A_E}+A_E}\right) - \mu_\varepsilon\, \varepsilon$. Note that we assume the basal rate of Fc$\varepsilon$RI in absence of IgE and alarmins is negligible, based on the fact that Fc$\varepsilon$RI complex is poorly expressed in non-atopic individuals[45]. Now, considering the dynamics of Fc$\varepsilon$RI at equilibrium (*i.e.* $\frac{d\varepsilon}{dt} = 0$) we get: $(\varepsilon)^\star = \frac{\rho_\varepsilon}{\mu_\varepsilon}(D)^\star\left(\frac{Y_A}{k_M^{Y_A}+Y_A} + \frac{A_E}{k_M^{A_E}+A_E}\right)$.

From this, we can set $v_{max}^{P_I} = f(\varepsilon) = (\varepsilon)^\star$, which we re-write as follows: $v_{max}^{P_I} = v_{max}^{\rho D}\left(\frac{Y_A}{k_M^{Y_A}+Y_A} + \frac{A_E}{k_M^{A_E}+A_E}\right)$, with $v_{max}^{\rho D} = \frac{\rho_\varepsilon}{\mu_\varepsilon}(D)^\star$.

Finally, we account for the down-regulation of antigen-presenting functions in presence TCS[47–52] (regulation $\mathcal{R}_-^{C,\rho D}$), which gives

$$\mathcal{R}_+^{\rho D} := \frac{v_{max}^{\rho D}\left(\frac{Y_A}{k_M^{Y_A}+Y_A} + \frac{A_E}{k_M^{A_E}+A_E}\right)(P_I)^2}{\left(k_M^{\rho D}\right)^2 + (P_I)^2}\left(1 + \mathcal{R}_-^{C,\rho D}\right) \tag{S12}$$

As IDECs display a higher surface expression of Fc$\varepsilon$RI than LCs[45], we used a scaling factor for the saturation constant of the antigen-dependent pathway of IDECs activation: $\mathcal{R}_+^{\rho D_E} = s_{D_E}\,\mathcal{R}_+^{\rho D} = s_{D_E}\,\mathcal{R}_+^{\rho D_L}$.

TYPE-1 ($Y_1$) grouping IL-12 and IFN-$\gamma$[18,43,45,46]

TYPE-2 ($Y_2$) grouping IL-4 and IL-13[27,45,46,55,66].

TYPE-17 & 22 with IL-17 ($Y_{17}$), IL-22 ($Y_{22}$), and S100A ($Y_S$) anti-microbial proteins[42,55,57]

REGULATORY ($Y_R$) grouping IL-10 and TGF-$\beta$[6,54]

We set the dynamics of any cytokine $X$ to follow a production (rate $\rho_X$) – decay (rate $\mu_X$) formalism. We further detail the modeling formalism of each functional group below.

#### Alarmins

Based on the litterature knowledge, we set the alarmins to be released by keratonocytes ($K$) only when the skin is damaged (term $(1-S)$)[37]. Furthermore, alarmin expression have been shown to be amplified by pro-inflammatory and type-2 cytokines[59,61,62], which we account for via the regulation $\mathcal{R}_+^K$.

$$\frac{\mathrm{d}Y_A}{\mathrm{d}t} = \rho_{Y_A}\, K\, (1-S)\, \left(1+\mathcal{R}_+^K\right) - \mu_{Y_A}\, Y_A \tag{S13}$$

#### Pro-inflammatory

Pro-inflammatory cytokines are released by DC[60,65], and this release would be amplified under antigen-dependent pathway of DC activation, via antigen uptake by Fc$\varepsilon$RI – IgE complex at the cell surface[43,45]. We account for this production term via the regulation $\mathcal{R}_{prod}^{\rho D}$ (Box S2). Note that this formalism allows to account for the fact that IDEC have been shown to release higher levels of pro-inflammatory cytokines than LC[43,45], which we assume to be linked to the observation that IDEC express higher levels of Fc$\varepsilon$RI (Box S2). TCS application have been shown to reduce, via various pathways, the release of pro-inflammatory cytokines[47,50–52,67,68], which is accounted for via the down-regulation $\mathcal{R}_-^{C,infl}$. Combining the different terms, the production of pro-inflammatory cytokines is then formalized as follows: $\mathcal{R}_{prod}^{C,\rho D} := D_L\, \left(1+\mathcal{R}_+^{\rho D}-\mathcal{R}_-^{C,infl}\right) + D_E\, \left(1+s_{D_L}^{D_E}\, \mathcal{R}_+^{\rho D}-\mathcal{R}_-^{C,infl}\right)$.

$$\frac{\mathrm{d}Y_I}{\mathrm{d}t} = \rho_{Y_I}\, \mathcal{R}_{prod}^{C,\rho D} - \mu_{Y_I}\, Y_I \tag{S14}$$

#### Type-1 cytokines

Type-1 cytokines are produced by Th-1 cells[43,45] under a type-1 cytokine environment[66,69]. This is implemented in our model following case-1 of adaptive response simplification (Box S3), via the term $\rho_{Y_1}\, D_E\, \mathcal{R}_+^{\rho D_E}$. As type-1 chemokines are chemo-attractants of Th-1 cells[66,69], we set the Th1-derived release of type-1 cytokines to be up-regulated by type-1 chemokines (regulation $\mathcal{R}_+^{Y_1,H_1}$). Type-1 cytokine synthesis is also assumed to be down-regulated by regulatory cytokines as they inhibit both type-1 and type-2 responses[2,18,54], which we account for in our model via the regulation $\mathcal{R}_-^{Y_R}$. Finlay, TCS application have been shown to limit the type-1 cytokine production by DC[49,50,70], which we implemented as for Pro-inflammatory via the regulation $\mathcal{R}_-^{C,infl}$.

We additionally account for type-1 cytokines production by innate memory-like cells ($D_M$) which are present under OM-85 treatment only.

$$\frac{\mathrm{d}Y_1}{\mathrm{d}t} = \rho_{Y_1}\, D_E\, \mathcal{R}_+^{\rho D_E} \left(1 + \mathcal{R}_+^{Y_1,H_1} + \mathcal{R}_-^{Y_R} - \mathcal{R}_-^{C,infl}\right) + \rho_{Y_1}^{D_M}\, D_M\, \mathcal{R}_+^{\rho D_E} \;-\; \mu_{Y_1}\, Y_1 \tag{S15}$$

**Type-2 cytokines**

Type-2 cytokines are released by type-2 Innate Lymphocyte Cells ($L$)[41,42], which we implemented in our model with the term $\rho_{Y_2}^{L}\, L$. Type-2 cytokines are also produced by Th-2 cells[27,43,45,46] under a type-2 cytokine environment[2,18,54]. This is implemented in our model following case-1 of adaptive response simplification (Box S3), via the term $\rho_{Y_2}^{D_L}\, D_L\, \mathcal{R}_+^{\rho D}$. As type-2 chemokines are chemo-attractants of Th-2 cells[66,71], we set the Th2-derived release of type-2 cytokines to be up-regulated by type-2 chemokines (regulation $\mathcal{R}_+^{Y_2,H_2}$). Type-2 cytokine synthesis is also assumed to be down-regulated by regulatory cytokines as they inhibit both type-1 and typ-2 responses[2,18,54], which we account for in our model via the regulation $\mathcal{R}_-^{Y_R}$. Finlay, TCS application have been shown to limit the type-2 cytokine production by DC[2,18,54], which we implemented as for Pro-inflammatory via the regulation $\mathcal{R}_-^{C,infl}$.

$$\frac{\mathrm{d}Y_2}{\mathrm{d}t} = \rho_{Y_2}^{L}\, L + \rho_{Y_2}^{D_L}\, D_L\, \mathcal{R}_+^{\rho D} \left(1 + \mathcal{R}_+^{Y_2,H_2} + \mathcal{R}_-^{Y_R} - \mathcal{R}_-^{C,infl}\right) - \mu_{Y_2}\, Y_2 \tag{S16}$$

**IL-17**

IL-17 are produced by Th-17 cells[18] in the presence of TGF-$\beta$, IL-1$\beta$, and IL-6[42,46,72]. This is implemented in our model following case-2 of adaptive response simplification (Box S3), via the term $\rho_{Y_{17}}^{T_{17}}\,(D_L + s_{D_L}^{D_E}\, D_E)\, \mathcal{R}_+^{\rho D}\, \mathcal{R}_+^{T_{17}}$. Furthermore, IL-17 production have been shown to be down-regulated by type-1 and type-2 cytokines[73], which we implemented in our model via the regulation $\mathcal{R}_-^{Y_{17}}$.

$$\frac{\mathrm{d}Y_{17}}{\mathrm{d}t} = \rho_{Y_{17}}^{T_{17}}\,(D_L + s_{D_L}^{D_E}\, D_E)\, \mathcal{R}_+^{\rho D}\, \mathcal{R}_+^{T_{17}}\, \mathcal{R}_-^{Y_{17}} - \mu_{Y_{17}}\, Y_{17} \tag{S17}$$

**IL-22**

As IL-17, IL-22 can be produced by Th-17[18] so we implemented the exact same production term than for IL-17: $\rho_{Y_{22}}^{T_{17}}\,(D_L + s_{D_L}^{D_E}\, D_E)\, \mathcal{R}_+^{\rho D}\, \mathcal{R}_+^{T_{17}}$. IL-22 can also by produced by Th-22[27,72] in the presence of TNF-$\alpha$ and IL-6[72]. This is implemented in our model following case-2 of adaptive response simplification (Box S3), via the term $\rho_{Y_{22}}^{T_{22}}\,(D_L + s_{D_L}^{D_E}\, D_E)\, \mathcal{R}_+^{\rho D}\, \mathcal{R}_+^{Y_{22}}$. We assume $\rho_{Y_{22}} := \rho_{Y_{22}}^{T_{17}} = \rho_{Y_{22}}^{T_{22}}$, such that IL-22 dynamics writes as follows: .

$$\frac{\mathrm{d}Y_{22}}{\mathrm{d}t} = \rho_{Y_{22}}\,(D_L + s_{D_L}^{D_E}\, D_E)\, \mathcal{R}_+^{\rho D}\, (\mathcal{R}_+^{T_{17}} + \mathcal{R}_+^{Y_{22}}) - \mu_{Y_{22}}\, Y_{22} \tag{S18}$$

**S100A**

S100A are released by keratinocytes ($K$) and this production have been shown to be amplified by IL-17 and IL-22[17,18,27], which we accounted for via the two regulations $\mathcal{R}_+^{Y_S,Y_{17}}$ and $\mathcal{R}_+^{Y_S,Y_{22}}$ respectively.

$$\frac{\mathrm{d}Y_S}{\mathrm{d}t} = \rho_{Y_S} \times K \times \left(1 + \mathcal{R}_+^{Y_S,Y_{17}}\right) \times \left(1 + \mathcal{R}_+^{Y_S,Y_{22}}\right) - \mu_{Y_S} \times Y_S \tag{S19}$$

**Regulatory cytokines**
Regulatory cytokines are released by naive DCs, which allows to maintain homeostasis[60]. In our model, we assume that only naive Lcs are involved in this pathway because (i) LCs represent the main epidermal DC sub-population in non-lesional skin and (ii) it has been suggested that a rapid influx of IDECs might be responsible for the breakdown of tolerance and onset of inflammation in AD patients[60]. As we do not explicitly represent the dynamics of naive LC, but assume a constant concentration $c_{D_L}$ (section S1.1.7), the release of regulatory cytokines by naive DC is formalized by the term $\rho_{Y_R}\, c_{D_L}$ in our model.

Regulatory cytokines are also released by regulatory T-cells ($T_R$)[54], so we implemented the following production term in our model: $\rho_{Y_R}\, T_R$.

Antigen uptake by DC (antigen-dependent pathway) can induce the release regulatory cytokines if DC express high level of Fc$\varepsilon$RI on their cell surface[42,43,45,46]. As IDECs display higher expression of Fc$\varepsilon$RI than LCs[45], we assume in our model that only IDECs can release regulatory cytokines following antigen uptake. The corresponding term is set as following: $\rho_{Y_R}\, D_E\, \mathcal{R}_+^{\rho D_E}$ (Box S2, Equation (S20)).

Finally, TCS application have been shown, via various pathway, to amplify the release of regulatory cytokines[50,52,68,70,74–77], which we accounted for via the regulation $\mathcal{R}_+^{C,Y_R}$.

$$\frac{\mathrm{d}Y_R}{\mathrm{d}t} = \rho_{Y_R}\,(1 + \mathcal{R}_+^{C,Y_R})\left(c_{D_L} + T_R + D_E\, \mathcal{R}_+^{\rho D_E}\right) - \mu_{Y_R}\, Y_R \tag{S20}$$

##### S1.1.12 Polyclonal IgE

**Biological mechanisms** Skin IgE consists of polyclonal IgE[81], released by IgE$^+$ plasmablasts in the skin tissues. Before infiltrating the skin tissues, IgE$^+$ plasmablasts are generated in the lymph nodes by IgE$^+$ B-cells[82]. IgE$^+$ B-cells differentiate from naive B-cells that bind consecutively to APC and helper T-cells (notably Th-2 cells[83])[80,84] under a type-2 cytokine environment[84,85]. Antigen presentation by APC to naive helper T-cells in the lymph nodes and their subsequent differentiation[78–80] is described in Box S3.

**Modeling formalism of IgE release** Very similarly to the simplification made for the release of the adaptive-derived cytokines (Box S3), we do not explicit the dynamics of IgE$^+$ B-cells neither of IgE$^+$ plamablasts in our model but represent IgE ($A_E$) release (rate $\rho_{A_E}$) by DC. Here we demonstrate that this simplification does not relies on strong assumptions.

From the detailed biological mechanisms, we can write the following ODE system to represent the synthesis of *IgE* antibodies, where $T_2$ are th-2 cells, $B$ IgE$^+$ B-cells, $P$ IgE$^+$ plasmoblasts, and $\mathcal{R}_{env}^{A_E}$ the level (between 0 and 1) of type-2 cytokine environment:

$$\begin{aligned}
\frac{\mathrm{d}B}{\mathrm{d}t} &= \alpha_B\, T_2\, \frac{D}{1+D} \mathcal{R}_{env}^{A_E} - \mu_B\, B \\
\frac{\mathrm{d}P}{\mathrm{d}t} &= \alpha_P\, B - \mu_P\, P \\
\frac{\mathrm{d}A_E}{\mathrm{d}t} &= \rho_{A_E}^{P}\, P - \mu_{A_E}\, A_E
\end{aligned}$$

From the modeling formalism of the adaptive response we adopted (Box S3) and under equilibrium for Th-2, B, and plasmablasts

**Box S3.** Modeling formalism of adaptive cytokine secretion

*Biological mechanisms* Once activated via an antigen-dependent pathway (1), DC express surface markers including antigens (2) and migrate towards the closest lymph nodes (3) where they present the antigen to naive CD4+ helper T cells[78–80]. Depending on the cytokine environment inside the LN, naive helper T-cells differentiate (4) towards a given type X of helper T-cells and then proliferate[80]. Finally, helper T-cells are attracted in the inflamed tissues (5) where they release specific cytokines (6).

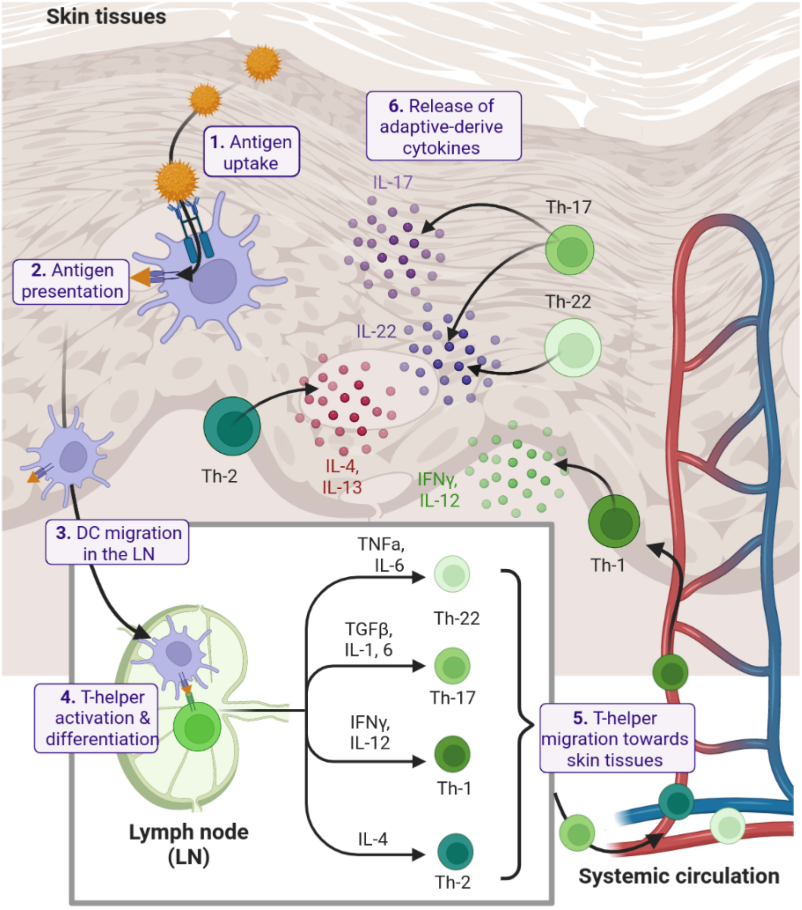

*Formalism* We do not explicit the dynamics of helper T-cells but we do account for the associated release of the adaptive derived cytokines, representing their synthesis by DC directly. As we are not interested in reproducing the kinetic of the immune response in details, this simplification is assume to have a negligible impact on the model behavior. We detail below how we can formalise adaptive-derived release of cytokines by DC, (A) starting from the detailed formalism, then (B) assuming helper T-cells at equilibrium (*i.e.* $\frac{dT_X}{dt} = 0$), and finally (C) using a condensed notation. For this simplification, two cases need to be distinguished:

**Case 1** Cytokines $Y_X$ released by DC $D_X$ induces the differentiation towards helper T-cells $T_X$, and helper T-cells $T_X$ release the same cytokines $Y_X$. This is the case of type-1 (Th-1) and type-2 (Th-2) helper T-cells.
**Case 2** Cytokines $Y_{X1}$ released by DC $D_X$ induces the differentiation towards helper T-cells $T_{X2}$, and helper T-cells $T_{X2}$ release the different cytokines $Y_{X2}$. This is the case of type-17 (Th-17) and type-22 (Th-22) helper T-cells.

| | **Case 1** | **Case 2** |
|---|---|---|
| (A) | $\begin{cases} \frac{dT_X}{dt} = \alpha_{T_X}\, \mathcal{R}_+^{\rho D}\, D_X - \mu_{T_X}\, T_X \\ \frac{dY_X}{dt} = \rho_{Y_X}^{T_X}\, T_X - \mu_{Y_X}\, Y_X \end{cases}$ | $\begin{cases} \frac{dT_{X2}}{dt} = \alpha_{T_{X2}}\, \mathcal{R}_+^{\rho D}\, D_X\, \mathcal{R}_+^{Y_{X1}} - \mu_{T_{X2}}\, T_{X2} \\ \frac{dY_{X2}}{dt} = \rho_{Y_{X2}}^{T_{X2}}\, T_{X2} - \mu_{Y_{X2}}\, Y_{X2} \end{cases}$ |
| (B) | $\begin{cases} (T_X)^\star = \frac{\alpha_{T_X}}{\mu_{T_X}}\, \mathcal{R}_+^{\rho D}\, D_X \\ \frac{dY_X}{dt} = \rho_{Y_X}^{T_X}\, (T_X)^\star - \mu_{Y_X}\, Y_X \end{cases}$ | $\begin{cases} (T_{X2})^\star = \frac{\alpha_{T_{X2}}}{\mu_{T_{X2}}}\, \mathcal{R}_+^{\rho D}\, D_X\, \mathcal{R}_+^{Y_{X1}} \\ \frac{dY_{X2}}{dt} = \rho_{Y_{X2}}^{T_{X2}}\, (T_{X2})^\star - \mu_{Y_{X2}}\, Y_{X2} \end{cases}$ |
| (C) | $\frac{dY_X}{dt} = \rho_{Y_X}\, D_X\, \mathcal{R}_+^{\rho D} - \mu_{Y_X}\, Y_X$ with $\rho_{Y_X} = \frac{\rho_{Y_X}^{T_X}\, \alpha_{T_X}}{\mu_{T_X}}$ | $\frac{dY_{X2}}{dt} = \rho_{Y_{X2}}\, \mathcal{R}_+^{\rho D}\, D_X\, \mathcal{R}_+^{Y_{X1}} - \mu_{Y_{X2}}\, Y_X$ with $\rho_{Y_{X2}} = \frac{\rho_{Y_{X2}}^{T_{X2}}\, \alpha_{T_{X2}}}{\mu_{T_{X2}}}$ |

we get:

$$(T_2)^\star = \frac{\alpha_{T_2}}{\mu_{T_2}} \mathcal{R}_+^{\rho D} D_L \quad \Leftarrow \begin{cases} (B)^\star &= \frac{\alpha_B}{\mu_B} (T_2)^\star \mathcal{R}_{env}^{A_E} \\ (P)^\star &= \frac{\alpha_P}{\mu_P} (B)^\star \\ \frac{\mathrm{d}A_E}{\mathrm{d}t} &= \rho_{A_E}^P (P)^\star - \mu_{A_E} A_E \end{cases}$$

From this, we can define $\rho_{A_E} = \rho_{A_E}^P \frac{\alpha_P}{\mu_P} \frac{\alpha_B}{\mu_B} \frac{\alpha_{T_2}}{\mu_{T_2}}$, which gives the following simplified dynamics of IgE, as implemented in our model:

$$\frac{\mathrm{d}A_E}{\mathrm{d}t} = \rho_{A_E} D_L \mathcal{R}_+^{\rho D} \mathcal{R}_{env}^{A_E} - \mu_{A_E} A_E \tag{S21}$$

###### S1.1.13 Polyclonal IgA

Mirroring the mechanisms of action of OM-85 in the context of respiratory tract infections[53], we assume the homing in the skin tissues of IgA$^+$ plasmablasts $P_{A_A}$ (which release polyclonal IgA $A_A$ in the skin tissues) from the systemic circulation under OM-85 treatment. Generation and trafficking of IgA$^+$ plasmablasts is set as in the already published model from Arsène *et al.* (2022)[53]. The only difference is their infiltration in skin tissues as well, but which is implemented in our model following the same formalism:

$$\begin{aligned} \frac{\mathrm{d}P_{A_A}}{\mathrm{d}t} &= \varphi\,(1-\sigma)\,P_{A_A}^{v} - \mu_{P_{A_A}} P_{A_A} \\ \frac{\mathrm{d}A_A}{\mathrm{d}t} &= \rho_{A_A} P_{A_A} - \mu_{A_A} A_A \end{aligned} \tag{S22}$$

###### S1.1.14 AD Severity

The model outputs AD severity using EASI (Eczema Area and Severity Index) and SCORAD (SCORing Atopic Dermatitis), which were identified as two of the three best validated outcome measures for AD severity[86].

- EASI score ($EASI$) is computed via the formula from Miyano et al. (2022)[87], which phenomenologically link AD severity with pathogen load ($P_I$) and skin barrier integrity ($S$). By definition, EASI varies between 0 and 72[86].

$$EASI = 72 \times \frac{P_I + (1-S)}{2} \tag{S23}$$

- SCORAD ($SCORAD$) is computed from EASI using a square function which has been fitted on literature data from Chopra *et al.* (2017)[88], Rullo *et al.* (2009)[89], and Yang *et al.* (2010)[90]. By definition, SCORAD varied between 0 and 103[91].

$$SCORAD = \sqrt{EASI \times 5.66 \times 10^{-3}} + 0.64 \tag{S24}$$

###### S1.1.15 Standard of care (SoC) administration

The administration of SoC ($SoC$, which represent either TCS: $C$ or emollients: $E$) is modeled by an on-off controller with a time step of one day. The choice to not explicit the pharmacokinetic of SoC relies on several assumptions:

- Once daily vs. twice daily applications have not been shown to give significantly different clinical results[92]. Therefore, it is reasonable to assume that neither intra-daily frequency nor total daily dose matters. Also, data linking dosage and clinical efficacy are too scarce for a correct calibration, as the dose applied is generally referred to as a "thin layer".

- The PK of topical corticosteroids can be disregarded, with a time to peak activity after administration of only two hours, and a penetration rate independent of skin state[50, 93–95].

- The PD of topical corticosteroids suggest a pharmacologic activity that lasts roughly one day[1, 96–99]. Adding an offset of six hours to account for the time to peak activity is arguably irrelevant with regard to the timescale of the simulation (weeks to months).

This time step still allows to simulate the two most commonly prescribed regimens: daily administration (acute/control phase) and twice weekly administration (maintenance/proactive phase).

#### S1.2 Model calibration

We here provide additional details on the procedure used for the model calibration. First, we detail how we constrained the disease model at equilibrium and the related assumptions we made (section S1.2.1). Then we present what data we used to inform the baseline biomakers and how we infer these baseline as functions of AD severity (section S1.2.2). Note that this was a crucial step to be able to constrain the disease model at equilibrium in treatment free context, but that the associated constrains can be relaxed and that baseline biomakers can be further inform with additional / new data set. Finally, we detail the calibration procedure of the model with aggregated data set on patient response to SoC (section S1.2.3).

##### S1.2.1 Constraints for model equilibrium assumption

We make the fundamental assumption that, without treatment, the disease state of a given patient characterized by a given disease severity is at equilibrium. This allows us to derive constraints on a number of parameters for the system to be at equilibrium. For this, we write the system with stationary variables: for all variables $X$, $\frac{\mathrm{d}X}{\mathrm{d}t} = 0$ and we note the level of a variable at equilibrium as $(X)^{\star}$.

The subset of variables considered as biomarkers (section S1.2.2) have, as such, their levels at equilibrium (or baseline) considered as input. Constraints for equilibrium can thus be written as functions of those baseline levels as well as other quantities. So that they can be used to inform parameter values, we make the following assumptions / choices:

- *Half-saturation constants* By definition, Hill functions $\mathcal{R}$ vary between 0 and the saturation constant ($v_{max}$), and reach half of its saturation value $\mathcal{R}_{50} := 0.5 \times v_{max}$ when the concentration of the modulator $X = k_M$. We assume that this mid-saturation point, $(\mathcal{R})^{\star}_{50}$ is reached at $EASI_{50} = 0.5 \times \max(EASI) = 36$. As a consequence, we set $k_M := (X)^{\star}_{50}$, the concentration (at equilibrium) of the biomaker(s) involved in the regulation at $EASI_{50}$. The only exceptions are:
  - The half-saturation constant of the up-regulation of immune cell activation by alarmin signals ($\mathcal{R}^{Y_A}_{+}$), which is calibrated.
  - The half-saturation constants of the up-regulation of antigen-dependent activation of DC ($\mathcal{R}^{\rho D}_{+}$), which are computed from the mid-point for skin barrier integrity ($(S)^{\star}_{50}$).

- *Mid-point for skin barrier integrity* As a fundamental hypothesis, we assume that the skin barrier integrity reaches half of it maximal value at $EASI_{50}$, *i.e.* $(S)^{\star}_{50} = 0.5$. As a consequence, the pathogen load ($P_I$) is constrained at 0.5 as well for an EASI of 36 (from Equation (S23)). Note that this hypothesis can be relaxed and was used to in deriving constrains for model equilibrium.

- *Saturation constants* $v_{max}$ are either calibrated or set to a constant.
- *Decay and degradation rates* $\mu$ are informed from the literature.

Finally, equilibrium constraints provide relationships between inputs (disease severity, biomarkers levels, half-saturation constants, saturation constants, informed parameters and calibrated parameters) and:

- skin barrier integrity $(S)^\star$ and pathogen infiltration load $(P_I)^\star$
- immune cells levels $(D_L)^\star$, $(D_E)^\star$, $(L)^\star$ and $(T_R)^\star$
- production rates $(\rho_X)^\star$ of biomarkers ($P_S$, $F$, $Y_S$, $Y_A$, $Y_I$, $Y_2$, $H_2$, $Y_1$, $Y_1$, $Y_{17}$, $Y_{22}$, $Y_R$)
- activation levels $(\alpha)^\star$
- carrying capacities $(c)^\star$

##### S1.2.2 Infer baseline biomakers

We infer the levels of selected biomarkers as a function of AD severity using literature data[55,100–103], the estimated relationships are provided in Tab. S4.

###### Immune system biomarkers

*Data selection, transformation, and extrapolation* There are numerous published data on skin levels of immune system biomarkers. In order to limit the bias due to the between protocol and population variability and uncertainty, we choose to focus on as few as possible data sources. As the primary target of this work is an AD pediatric population, we privileged data from children. As the model describes the dynamics of biomakers in the skin tissues at the between-cell scale, we privileged data from skin sampling quantifying biomaker levels rather than expression. We selected Lyubchenko *et al.* (2021)[101] as primary data source, which reports the levels of 7/11 immune biomarkers in our model from tape-stripping on a pediatric population. To complement the primary data source, we also consider two additional sources: Guttman-Yassky *et al.* (2019)[104] and He *et al.* (2020)[103], which report protein expression levels from tape-stripping.

In the primary data source (Lyubchenko *et al.* (2021) [101, Fig. 3]), biomarker levels are expressed as pg (biomarkers) / µg (total protein), the amount of total protein assumed to be the same for all patients and set to 150 µg/mL [101, Fig. 2]. Based on each biomarker LoQ from the MSD U-Plex human cytokine multiplex platform (MesoScaleDiagnostics, Rockville, Marylan), we assumed that biomarker data from Lyubchenko *et al.* (2021)[101] are exploitable if the median value is higher than a LoQ = 50 pg/mL (5 times the highest LoQ estimated from the MSD U-Plex platform). Corresponding transformed data are provided in Fig. S2. When biomakers where not informed in Lyubchenko *et al.* (2021)[101] or had a median levels below the LoQ, biomarker expression data (quantified with RT-PCR from skin tape strips) were used to complement the data (He *et al.* (2020) [103, Fig.2, 3, 5] and Guttman-Yassky *et al.* (2019) [104, Fig. 1B, 2A]). We assume that summary data of levels of biomarkers of the same functional group are correlated and can can be summed. Below are the detailed assumptions made based on the available data, for each functional category considered in our model (Tab. S5):

- **Alarmins** (Tab. S5[$Y_A$]): sum of levels of TSLP & IL-33 from Lyubchenko *et al.* (2021)[101], no data was found for IL-25, which is assumed to have negligible levels in the skin tissues. The median of the sum remains below the LoQ.
- **Pro-inflammatory cytokines** (Tab. S5[$Y_I$]): sum of levels of IL-8, IL1-$\beta$, and IL-6 from Lyubchenko *et al.* (2021)[101],

no data was found for TNF-$\alpha$, which is assumed to have negligible levels in the skin tissues.

- **Type-2 cytokines** (Tab. S5[$Y_2$]): IL-13 levels from Lyubchenko *et al.* (2021)[101], no data was found for IL-4. Overall, the median is assumed to be below the LoQ as Guttman-Yassky *et al.* (2019) reports IL-4 RT-PCR data lower than IL-13, and IL-13 levels from Lyubchenko *et al.* (2021)[101] are below the LoQ.

- **Type-2 chemokines** (Tab. S5[$H_2$]): sum of levels of CCL-17 & CCL-22 from Lyubchenko *et al.* (2021)[101], no data was found for CCL-27, which is assumed to have negligible levels in the skin tissues.

- **Type-1 cytokines** (Tab. S5[$Y_1$]): IL-12 levels from Lyubchenko *et al.* (2021)[101], no data was found for IFN-$\gamma$. Overall, the median is assumed to be below the LoQ as both Guttman-Yassky *et al.* (2019) and He *et al.* (2020) report IFN-$\gamma$ RT-PCR data lower than IL-13, and IL-13 levels from Lyubchenko *et al.* (2021)[101] are below the LoQ.

- **Type-1 chemokines** (Tab. S5[$H_1$]): no data was found for CXCL10, but assumed to be below the LoQ as both Guttman-Yassky *et al.* (2019) and He *et al.* (2020) report CXCL10 RT-PCR data similar than IL-13, and IL-13 levels from Lyubchenko *et al.* (2021)[101] are below the LoQ.

- **IL-17 & IL-22** (Tab. S5[$Y_{17}, Y_{22}$]): from Lyubchenko *et al.* (2021)[101], median below the LoQ.

- **Antimicrobial proteins S100A** (Tab. S5[$Y_S$]): no data were found. He *et al.* (2020) reports S100A7 and S100A8 RT-PCR data lower than S100A9, and both Guttman-Yassky *et al.* (2019) and He *et al.* (2020) report S100A12 RT-PCR data strongly lower than S100A9, while Guttman-Yassky *et al.* (2019) reports S100A9 RT-PCR data similar to IL-8. As a consequence, we assumed S100A summary levels equal to IL-8 from Lyubchenko *et al.* (2021)[101].

- **Regulatory cytokines** (Tab. S5[$Y_R$]): IL-10 levels from Lyubchenko *et al.* (2021)[101], no data was found for TGF-$\beta$, which is assumed to have negligible levels in the skin tissues. The median of the sum remains below the LoQ.

- **IgE** (Tab. S5[$A_E$]): no data was found, levels of IgE are assumed similar than alarmins (*i.e.* lower than the LoQ) such that both have a balanced effect on the up-regulation of the expression of Fc$\varepsilon$R receptor, involved in the antigen-dependent activation of dendritic cells (regulation defined in Eq. (S14)).

*Procedure* We assume there is a strong positive correlation between any biomarker levels and AD severity, based on tape stripping immunoassay data [101, Fig. 4] and RT-PCR expression level quantification from He *et al.* (2020) [103, Fig. 5] and Guttman-Yassky *et al.* (2019) [104, Tab. 2], assuming that trends observed in RT-PCR data can be translated on tape stripping immunoassay data as suggested by Lyubchenko et al. (2021) [101, Discussion]. For any immune biomarker, an exponential regression is fitted between biomarker data (median for healthy control: $X_{HC}$, and Q1, median, Q3 of values for AD patients) and the SCORAD values (Q1, median, Q3 respectively): $X = e^{\alpha_X \times SCORAD + log10(X_{HC})}$ as illustrated on Fig. S3.A. The resulting $\alpha_X$ values are provided in Tab. S4. To account the between-patients variability, a white noise $\sigma_X \sim \mathcal{N}(0, 1.25)$ is added as follows: $X = e^{\alpha_X \times SCORAD + \log 10(X_{HC}) + \sigma_X}$.

**Filaggrin** Contrary to the immune systems biomarkers we found only few studies focusing on filaggrin. We used data from Trzeciak *et al.* (2016) [105, Fig. 3] where the authors performed skin biopsies on a cohort of 19 moderate to severe AD patients (SCORAD = 62.2 ± 14.2) and 26 healthy subjects and measured filaggrin protein level as ng per g of wet tissue. We converted these values to ng/mL using a conversion factor of 1.1 g/mL, obtained from aquacalc. The authors do not provide the correspondence between AD severity and filaggrin values, but a strong negative correlation (-0.886) between both[105]. Based on this and extrapolated data, we obtained the following data table. Finnaly, similarly than the procedure used for the immune biomakers we here fitted a negative linear relationship between the log of filaggrin levels and the scorad (Tab. S4). The between-patient variability (Tab. S4) was set to best fit the boxplot of filaggrin levels in [105, Fig. 3], as illustrated in Fig. S3.B.

***S. aureus* colonization of the skin microbiome** Direct relationship between the % *S. aureus* and AD severity is not readily available in the literature. We compiled data from Sun *et al.* (2009)[106] Leung *et al.* (2019)[107]. While Leung *et al.* (2019)[107] provides individual data of the % of *S. aureus* alongside with the AD severity there is relatively limited samples with % *S. aureus* above 5 %. Sun *et al.* (2009) does not give a direct comparison of % *S. aureus* and SCORAD, however we were able to estimate of the % of *S. aureus* as a function of AD severity with the microbiome index of skin health (MISH) as intermediary (Fig. S3.C). Compiling both data we fitted a direct relationship between % of *S. aureus* and SCORAD (Fig. S3.C).

##### S1.2.3 Patient response to standards of care

The calibration objective was to reproduce the quantitative behavior of treatment efficacy (*i.e.* AD severity improvement percentage with respect to the baseline) of the standards of care (emollient and topical corticosteroids – mild and high potency). Five data sets from the literature have been used for the calibration of patient response to standard of care, corresponding to a pediatric population with a wide range of AD severity (Tab. S6).

**Calibration method** Our calibration procedure has been published elsewhere[108]. In brief we use global optimization as implemented in the covariance matrix adaptation evolution strategy (CMA-ES)[109] algorithm. A population size of 15 patients has been chosen. The objective optimization function is a modular scoring system suitable for data normalization[108], and which is evaluated across the 11 *in silico* trial arms used as scenarios. Parameters were initially sampled around 8 points in parameter hyperspace using latin hypercube sampling (multistarts) using an initial standard deviation of 0.2. All treatments are applied at 30 day of simulation and the observation frequency (tstep) is fixed to 1 day. The total simulation time depends on the scenario arm and matches the duration of the trials described by each data source. The algorithm was run for 100 iterations.

**Data** All selected datasets have been homogenized to use the relative improvement percentage, even when the absolute EASI measurements are available. Moreover, all datasets, where clinical outputs have been reported as SCORAD, have been converted into EASI (using a conversion function, see Section S1.1.14 and Equation (S23)). As there is high uncertainty in the severity score measurement, and due to lack of variability data, we deemed that using relative improvement percentage was the best way to proceed with the heterogeneous datasets. Narrow and wide bounds was arbitrary set up to reference value $\pm 1\%$ and reference $\pm 5\%$ respectively.

**Optimization criteria** We optimized the percentage of EASI improvement of the reference patient under SoC treatment. The optimization is based on five time series data sources and one source containing one time point (Tab. S6), resulting in six goodness-of-fit values. The selected reference patient at the end of the calibration procedure was the one with the best mean goodness-of-fit once the optimization algorithm converged. Goodness-of-fit values are between 0 and 1, with 1 corresponding to a perfect fit.

**Goodness of fit** The calibration is acceptable as the result correctly produces most of the treatment efficacy data (see Fig. S4), with a mean goodness-of-fit of 0.998. The lowest goodness of fit was of 0.922 (see Fig. S4, middle-left panel), for the time series data from Kim et al. (2020). Convergence was reached by the optimization algorithm. Parameter values associated with the best patient from the calibration procedure are provided in Tab. S7.

#### S1.3 Selection of biomakers

For optimal alignment of our mechanistic model with biomarker programs, we define a set of 13 variables represented in the model as biomarkers († mark in Tab. S1), inspired by existing knowledge on AD biomarkers. A biomarker is a measurable entity, predictive of response to treatment or disease severity and should be biologically relevant as well as linked to disease mechanisms[55]. We selected eleven immune biomarkers to cover a wide spectrum of AD endo/phenotypes: alarmin signals (TLSP, IL-33, IL-25); pro-inflammatory (IL-1$\beta$, IL-6, IL-8, TNF-$\alpha$); type-1 (chemokines: CXCL10 and cytokines: IL-12, IFN-$\gamma$); type-2 (chemokines: CCL-17, CCL-22, CCL-27; cytokines: IL-4, IL-13; antibodies: polyclonal IgE); type-17 and -22 (IL-17, IL-22, and S100A proteins); and regulatory (IL-10, TGF-$\beta$) responses. Indeed, there is a wide spectrum of AD endo/phenotypes that exhibit different levels of activation of the innate pro-inflammatory, type-1, regulatory, and type-17/22 responses[2,6,42], while independently of the phenotype, the immune response to AD exhibits a type-2 skewing[2,3,6,27,42,43]. We additionally select fillagrin level as marker of skin barrier integrity, and the colonization percentage by *S. aureus* in the skin microbiome.

#### S1.4 Boostrapping procedure to estimate sample size and power

Let X and Y the change in SCORAD in the placebo and treated groups respectively. For a given sample size n = 85, the power is computed by the following steps, applied for every bootstrap sample:

- Sample group G0 with n patients is drawn from X
- Sample group G1 with n patients is, independently from G0 drawn from X
- Sample group G2 with n patients is drawn from Y
- The mean of G0, G1, and G2 is computed
- The difference between G0 and G1 is computed to get a sample m0 following the H0 hypothesis
- The difference between G0 and G2 is computed to get a sample m1 following the H1 hypothesis

Samples of m0 are then used to derive the empirical distribution of the Student's t test under H0, while samples of m1 are used to derive the empirical distribution of the metric under H1. These empirical distributions are then used to find values where H0 is rejected for a confidence level  and compute the power of H1. This approach is repeated 1000 times, for sample sizes ranging from 10 to 300. The average power over the 1000 repetitions is computed. A spline function is then used to compute the exact sample size required to reach a given level of statistical power. The whole procedure is repeated 100 times, from which we extract the mean and standard deviation for both power and sample size.

### S2 Supplementary Results

#### S2.1 Assessment of the efficacy sensitivity to the TCS related trial protocol settings

We assessed the sensitivity of clinical trial efficacy to the duration of TCS induction phase, TCS potency, and TCS administration frequency via three metrics: mean clinical efficacy, sample size and power (Fig. S5). Our results showed that efficacy is not sensitive to any TCS related trial protocol settings (Fig. S5). We further investigated the model behaviour depending on the duration of TCS induction phase and TCS administration frequency.

##### S2.1.1 Exploration of the impact of TCS induction phase

Our results showed no impact of the TCS induction phase duration (IPD) on the treatment effect at 6 months, neither at the population scale (Fig. S5 A – C) nor at the individual scale with a reference patient (Fig. S6). However, there is an impact (at least for some patients) of the TCS IPD on efficacy at time points between 15 days and 2 months. In particular, the time at which the effect plateau is reached depends on the IPD and more importantly, whether longer IPD leads to shorter or longer time to effect plateau depends on considered arm (Fig. S6.A-B). In the treatment arm, longer induction phases lead to shorter time to maximum effect (Fig. S6.A), whereas it is the opposite in the placebo arm where shorter IPD leads to shorter time to maximum effect (Fig. S6.B). Indeed, in the placebo arm, since the IP includes a phase of TSC administration every day and since TSC are administered every 3 days once the IP is over, prolonged IPD results in a lower severity reached during the IPD than at the end of the trial. As a consequence, there is no impact of IPD on treatment effect which is a measure taking into accounts both arms (Fig. S6.C). Importantly, these conclusions might be patient-dependent. Indeed, our exploration demonstrated a high between-patient variability of the impact of IPD, as illustrated in Fig. S6.D – F. Comparing two patients with highly different AD endotypes (X and Y), it can be observed that the IDP has no impact on the treatment effect for patient X, and this at any point in time (Fig. S6.D) whereas for patient Y, the IDP has a strong impact on treatment effect for patient Y if measured before month 6 (e.g. at 3 months, vertical lines in Fig. S6.E).

##### S2.1.2 Exploration of the impact of TCS administration frequency

While TCS administration frequency does have an impact on the reference patient response (Fig. S7.A), there is no trend at the population scale (Fig. S7.B). To try to understand where this was coming from, we first assessed if the impact of TCS administration frequency was dependent on baseline AD severity since we observed that its impact is negligible for reference patients with high or low baseline SCORAD (30 vs reference = 40 vs 60, Fig. S7.C and D). To do so, we looked at the distribution of treatment effect under various scenarios of TCS administration frequency depending on SCORAD baseline (Fig. S7.C). Our results shows that there is no notable differences in the impact of the TCS administration frequency depending on the baseline scorad (Fig. S7.E and F). However, there is a clear positive trend between the treatment effect and the baseline scorad whatever the TSC administration frequency (Fig. S7.E and F). Note that while the impact of TCS administration frequency on the patient response seems to be negligible at the population scale, it can be marked at the patient level, depending on the patient characteristics. As the between-patient variability in the strength of the impact can not be attributed to the baseline SCORAD (Fig. S7.E and F), we can assume that this is due to the AD endotype variability (i.e. variability in baseline biomarkers). This hints that there may be relevant advanced cross-optimization strategy to consider where patients are selected based on whether they would benefit the most from an increased or decrease TCS administration frequency.

#### S2.2 Performance of baseline AD severity as a marker of response

Here, we assess to what extent the most simple biomarker, the patient's baseline AD severity, is predictive of OM-85 effect and more importantly if it can be used as a selection criteria for trial optimization. For this, we generated a virtual population where baseline AD severity fully determines the patient's biomarkers levels (Section S1.2.2, Tab. S4). A visual check and the coefficient of determination ($R^2 = 0.35$) indicates the poor performance of baseline severity as predictor of treatment effect (Fig. S8.A). In particular, similar predictions with the set of selected biomarkers (Fig.4) were a lot more accurate (Fig. S8.A). We then verified that the relative low performances of severity as a marker of treatment effect is not dependant on which severity range we are considering (Fig. S8.B). Finally, we assessed the potential for clinical trial optimization by selecting best responders predicted using the baseline severity only (Fig. S8.C). While our results showed a potential for increasing the average treatment effect and decreasing the sample size (as well as for increasing the proportion of successful trials, results not shown), the recruitment effort increases exponentially with the restrictiveness of the selection criteria (Fig. S8.A to C respectively). This suggests that the absence of selection is optimal and the low potential of baseline severity as a selection criteria for trial optimization.

#### S2.3 Identification of predictive biomarkers using a surrogate model approach

Complementary to the reduced virtual population approach presented in the main text, we also tested to identify markers predictive of treatment response using a more standard statistical approach. For this, the input-output transformation of the complete ODE model was approximated by a generalized linear model similar to what is often implemented in covariate efficacy analyses of clinical trials (Fig. S9.A). The linear model predicting the treatment effect from the baseline of the thirteen biomarkers was associated with a poor accuracy of prediction ($R^2 = 0.21$), which demonstrate the complexity of the mechanisms involved. To proceed with a complete statistical analysis approach, we then created all possible linear models using a step-wise method, including all the possible combination of one to thirteen biomarkers (Fig. S9.A). The prediction from each possible linear model was evaluated with the Bayesian Information Criterion (BIC, Fig. S9.A), which penalizes the number of biomakers included in the linear model. This has been achieved using the `betsglm` R package. We selected the linear model associated with the lowest BIC, which in this case included nine over the thirteen biomarkers (BIC, Fig. S9.B). We also evaluated the predictions of this optimal model by plotting the predicted treatment effect against the treatment effect of the corresponding patients from the fully informed virtual population (Fig. S9.D). As expected, this was associated with a low coefficient of determination ($R^2 = 0.20$). While this statistical analysis method was unreliable to select a set of predictive biomakers, the fourth-top of the most predictive biomakers was in agremment with the results from the alternative method presented in the main text (see Fig. S9.C vs Fig. 4 in the main text).

We additionally assessed the qualitative (positive or negative) link between each of the thirteen biomakers and the treatment effect: Fig. S9.C displays the sign of regression coefficients for each biomarker and for each of the best (*i.e.* with the lowest BIC) linear model including one to thirteen biomarkers. Finally, we evaluated the potential for realistic clinical trial optimization by selecting predicted responders with the optimal linear model (Fig. S10). While our results showed a potential decrease of the sample size (Fig. S10.B) and increase of the proportion of successful trials (Fig. S10.C), the potential for increasing the avaerage treatment effect was moderated (Fig. S10.A) and the recruitment effort increased exponentially with the restrictiveness of the selection criteria (Fig. S10.D). This suggests that the absence of selection is optimal.

### S3 Supplementary Figures

**Figure S1.** Graphical illustration of the links between the OM-85 induced GALT stimulation and the mechanisms of action in the lesional AD skin, as represented in our model.

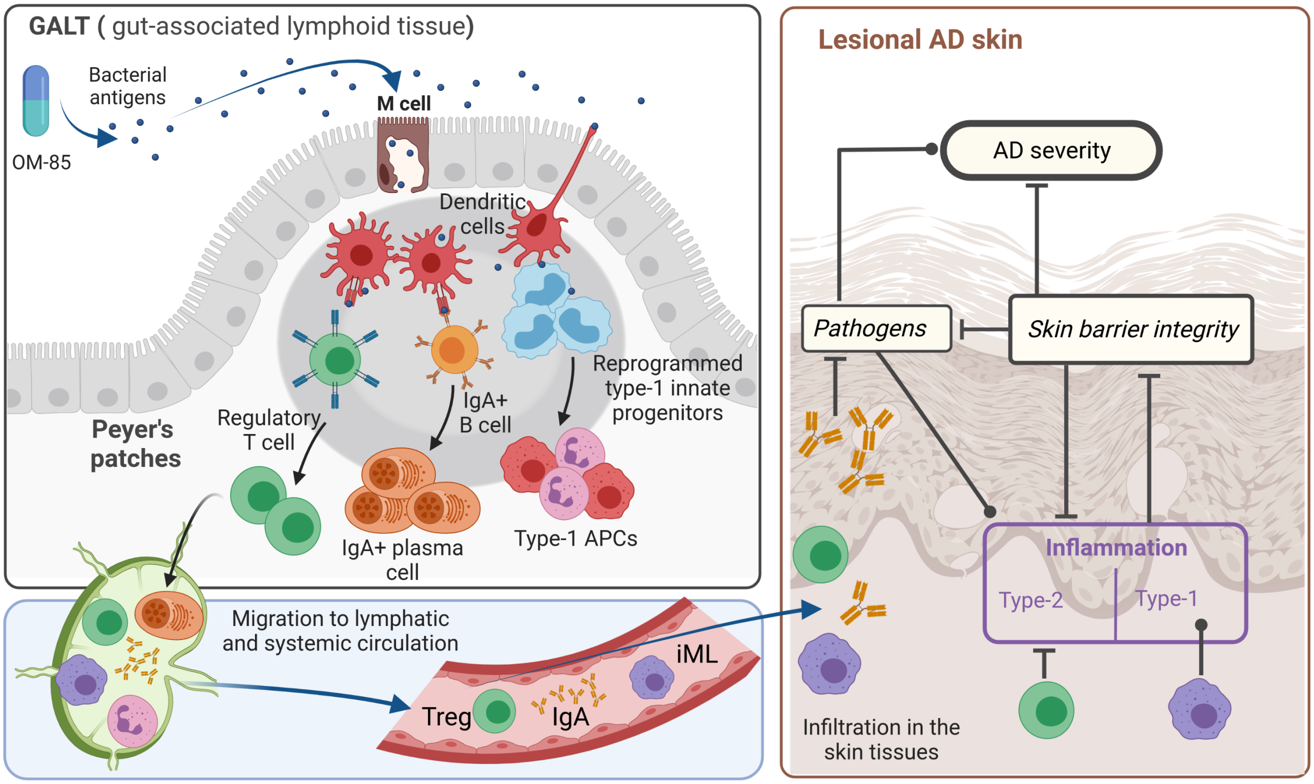

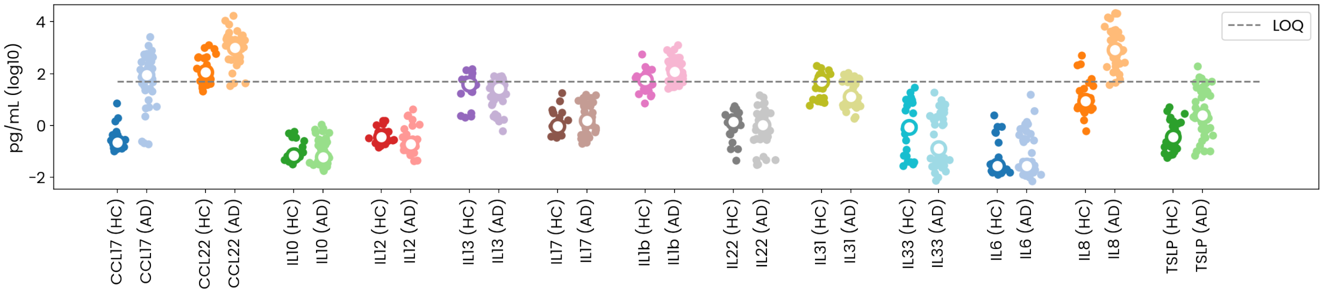

**Figure S2.** Absolute concentrations (pg/mL, log10 transformed) for immune biomarkers of interest for AD and healthy (HC) children (points: individual data, circle: median). Data was extracted from Lyubchenko *et al.* (2020)[101], a mean total protein concentration in the tape-strips extracts of 150 $\mu$g/mL was assumed based on values reported in the study to convert the reported pg/$\mu$g values to absolute concentrations. The Limit of Quantification (LOQ) was estimated at 50 pg/mL from MSD U-Plex human cytokine multiplex platform (MesoScale Di-agnostics, Rockville, Maryland) and is indicated with a dash grey line.

**Figure S3. Estimation of the relationship between biomakers and AD severity**

**A. immune biomakers** (illustration)

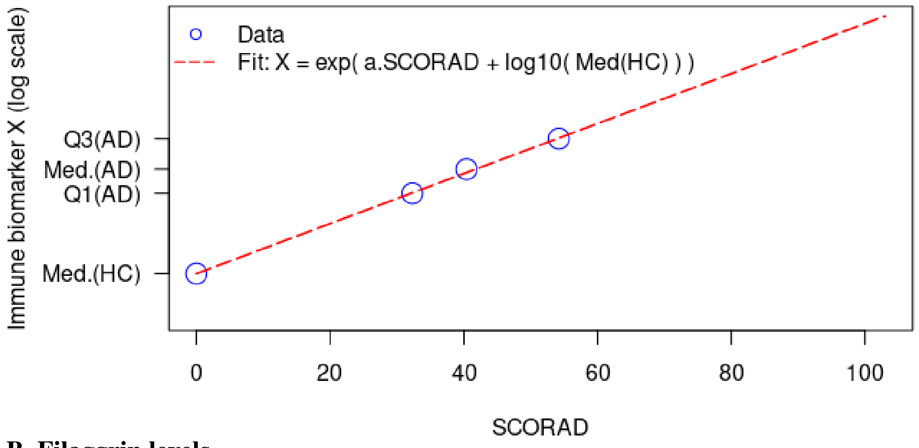

**B. Filaggrin levels**

Comparison of estimated levels from the fit with the original figure [105, Fig. 3]

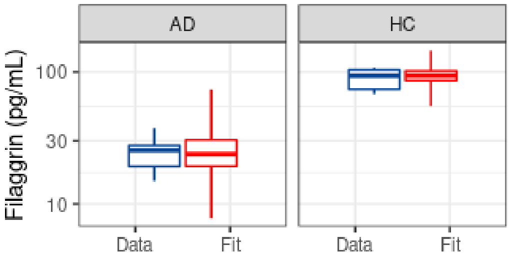

Extrapolated data from Trzeciak *et al.* (2016) [105, Fig. 3]:

| | Healthy Median | AD patients Q1 | AD patients Median | AD patients Q3 |
|---|---|---|---|---|
| SCORAD | 0 | 52.6 | 62.2 | 71.7 |
| Fillagrin levels (pg/mL) | 93.9 | 27.8 | 25.5 | 19.2 |

**C: % *S. aureus*.** 1. Estimate of the relationship between the % of *S. aureus* and the microbiome index of skin health (MISH). 2. MISH depending on SCORAD (data in Sun *et al.*), from which, combined with 1., estimates of the percentage of *S. aureus* depending on AD severity are computed. 3. Fitting of the relationship between the % of *S. aureus* and AD severity, compiling estimates from Sun *et al.* (2009) and data from Leung *et al.* (2019).

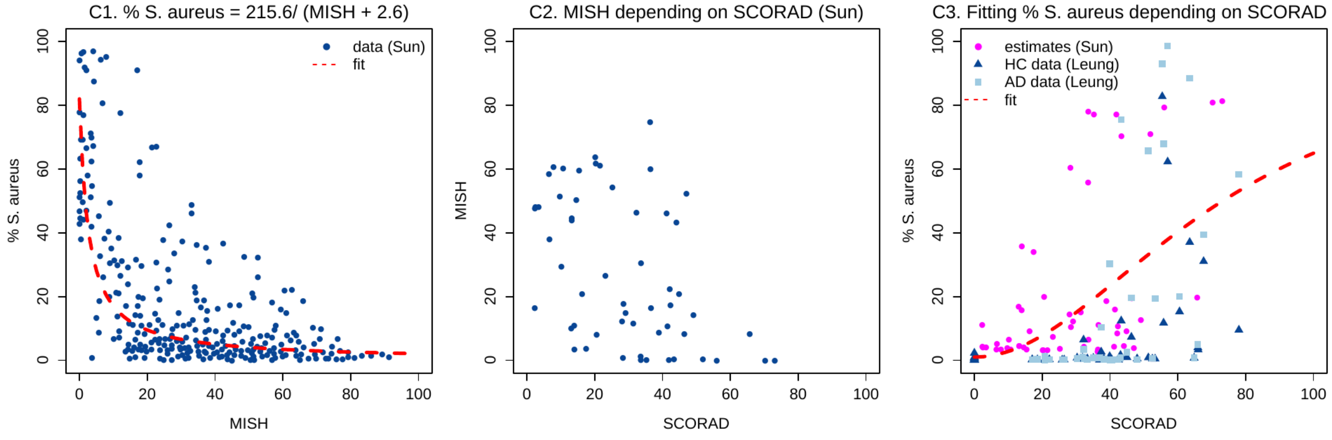

**Figure S4.** Best patient from SoC calibration: comparison with calibration data and associated goodness-of-fit (between 0 and 1 by definition, where 1 corresponds to a perfect fit). In the simulation, treatments start at day 30, which corresponds to day 0 of the data. The first time point in each panel corresponds to the first time point of the corresponding data set

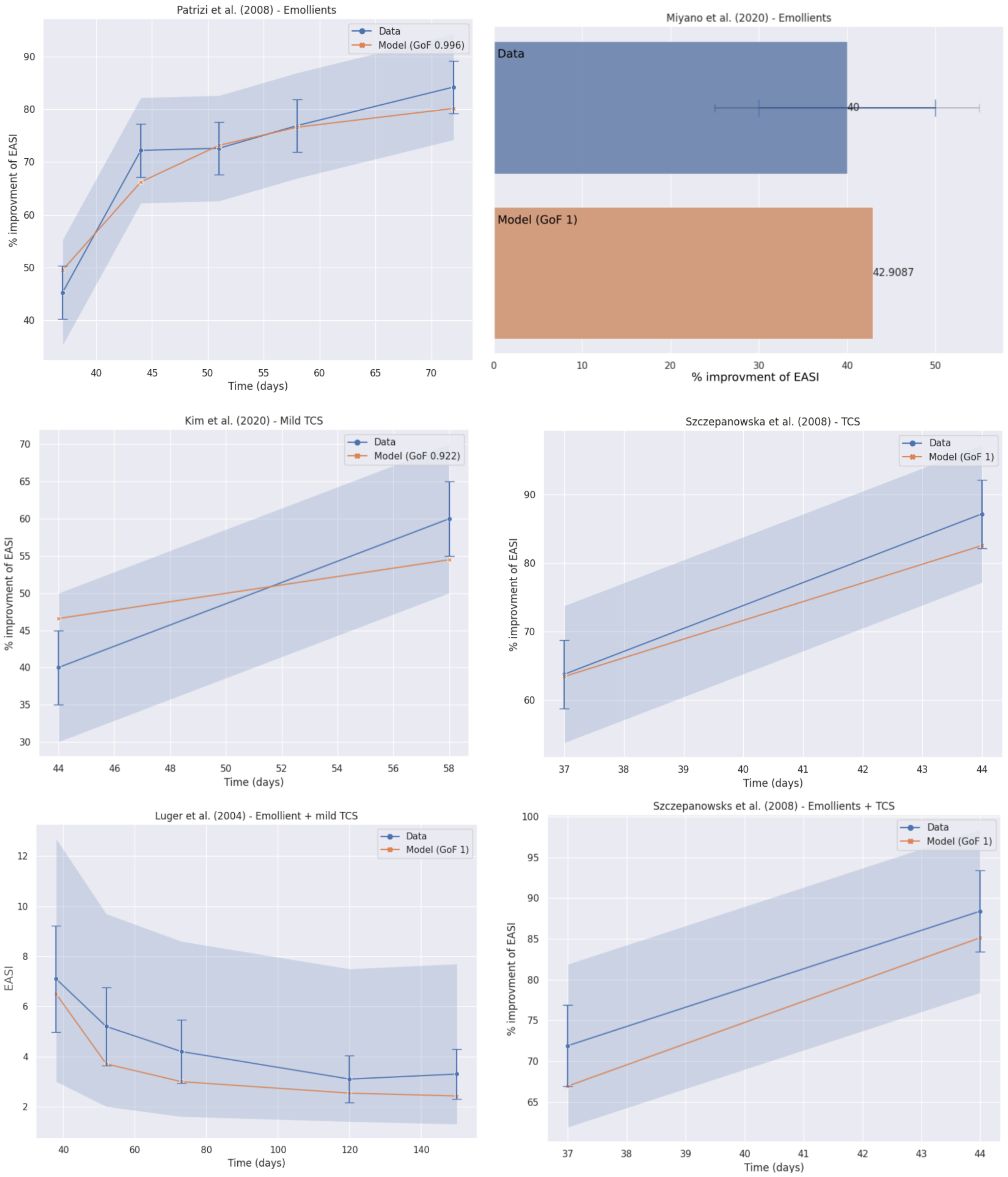

**Figure S5. Sensitivity of the clinical trial efficacy** to duration of the induction phase (A – C), the TCS potency (D – F), and the TCS administration frequency (G – I). Clinical trial efficacy is quantified via three metrics: **mean clinical efficacy** (A, C, D: median – line, interquartile range – box, whisker – dashed line); **sample size distribution** (B, E, H: obtained by bootstrapping over 100 samples, mean – line and standard deviation – colored area); **statistical power** (i.e. proportion of successful trials, C, F, I: obtained by bootstrapping over 100 samples, mean – line and standard deviation – colored area).

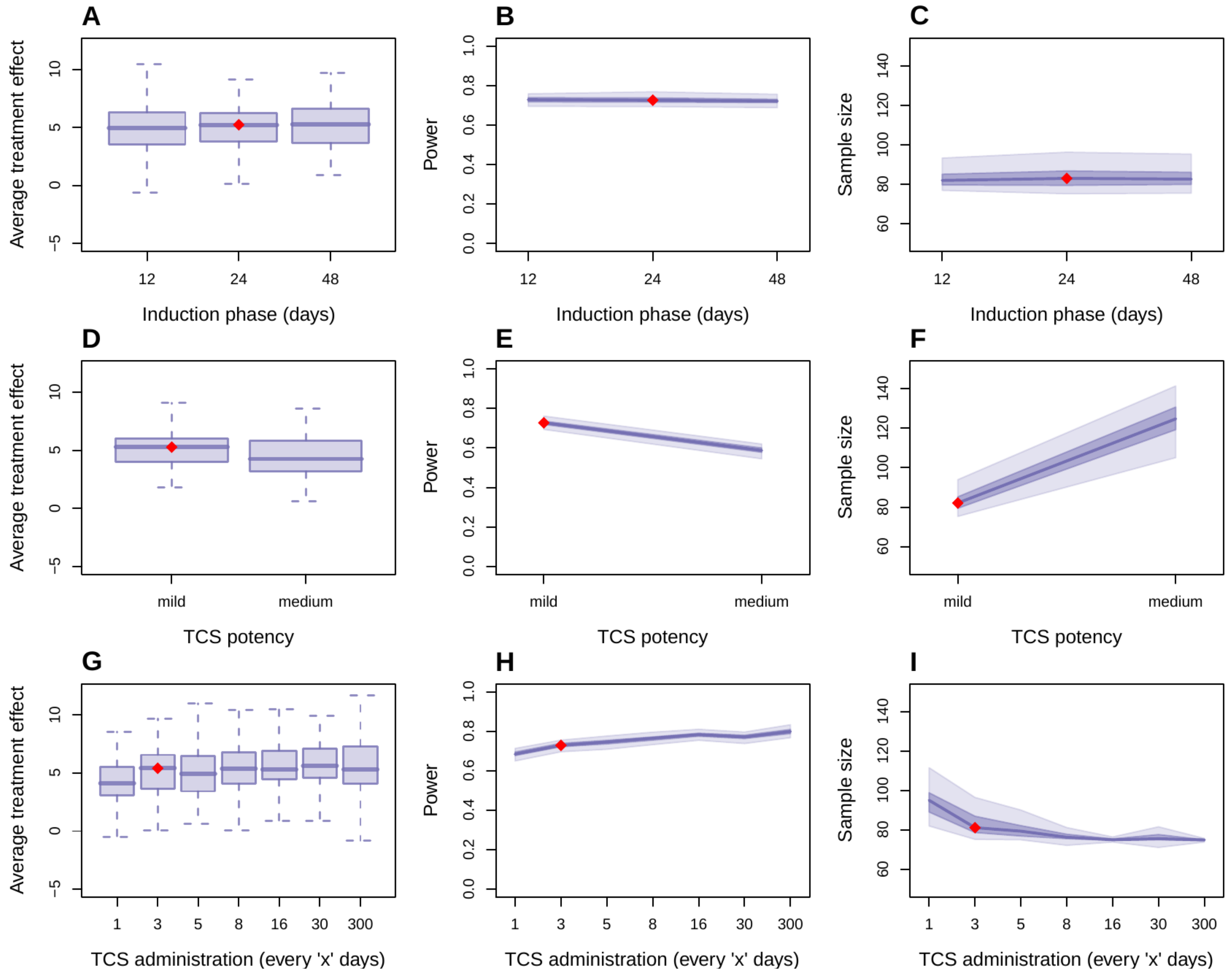

**Figure S6.** Exploration of the potential impact of TCS induction phase duration (IPD) on the clinical efficacy. Clinical efficacy is given by the difference in the SCOARD between the placebo and treatment arm for a given patient. **A – C** Severity evolution for the reference patient depending on induction phase duration (IDP) for A treatment arm, B placebo arm, and C treatment vs placebo arms for the two extrem IDP (no: no induction phase, sp: super prolonged). **D – F** Clinical efficacy (treatment vs placebo arms) efficacy depending on the IDP for two random patients X (D) and Y (E) and X vs Y patient (F).

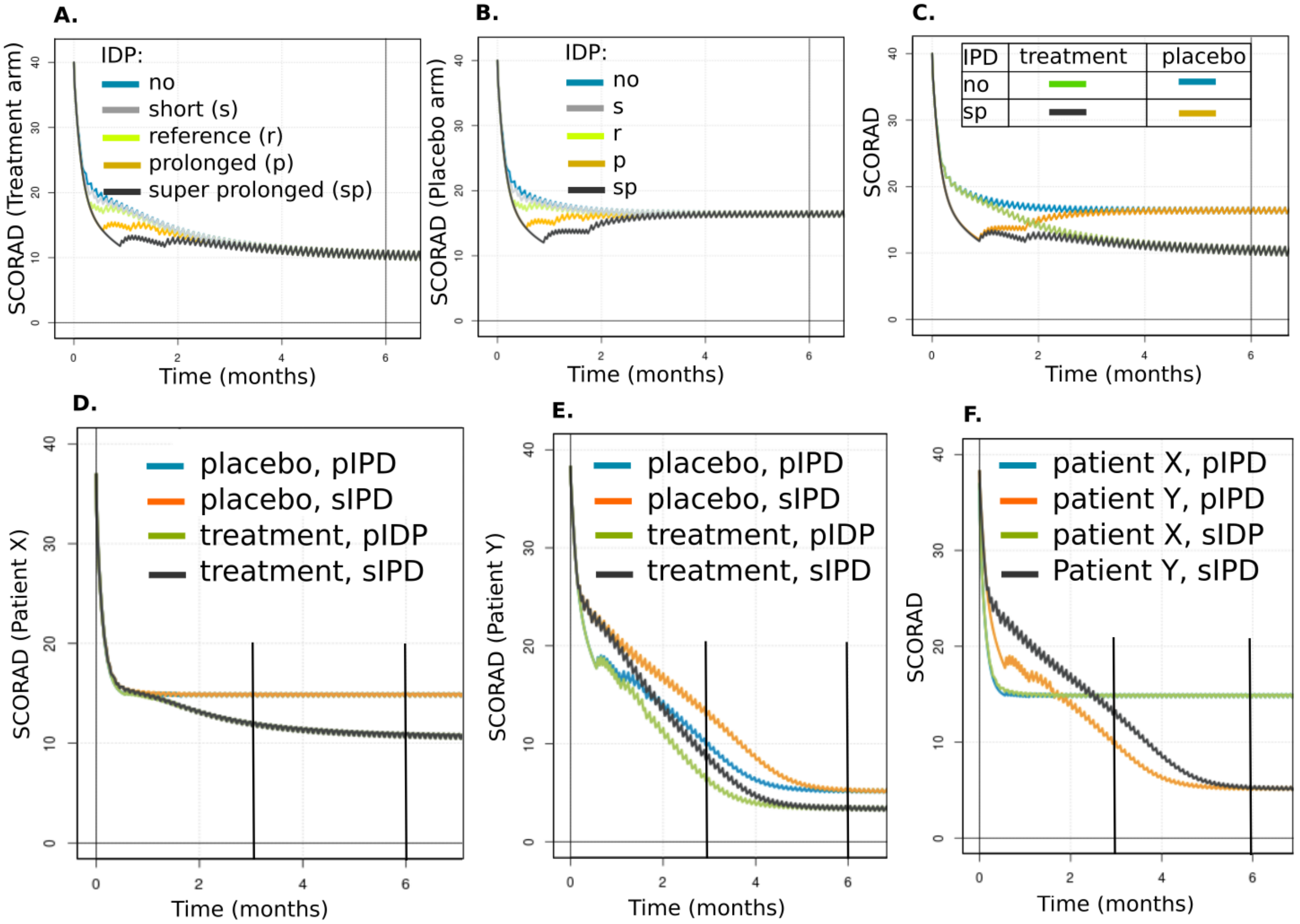

**Figure S7.** Exploration of the potential impact of TCS frequency administration on the clinical efficacy. **A, C, D** Clinical efficacy (SCORAD for placebo vs treatment arms) evolution for the reference patient depending on TCS administration frequency and SCORAD baseline (A. 40, C. 30, D. 60). **B, E, F** Distribution of clinical efficacy over the virtual population depending on **B.** TCS administration frequency from every day (e1D) to every 300 days (e300D) or depending on baseline SCORAD and **E.** without TCS administration (∼ e300D), **F.** TCS administration every day (e1D).

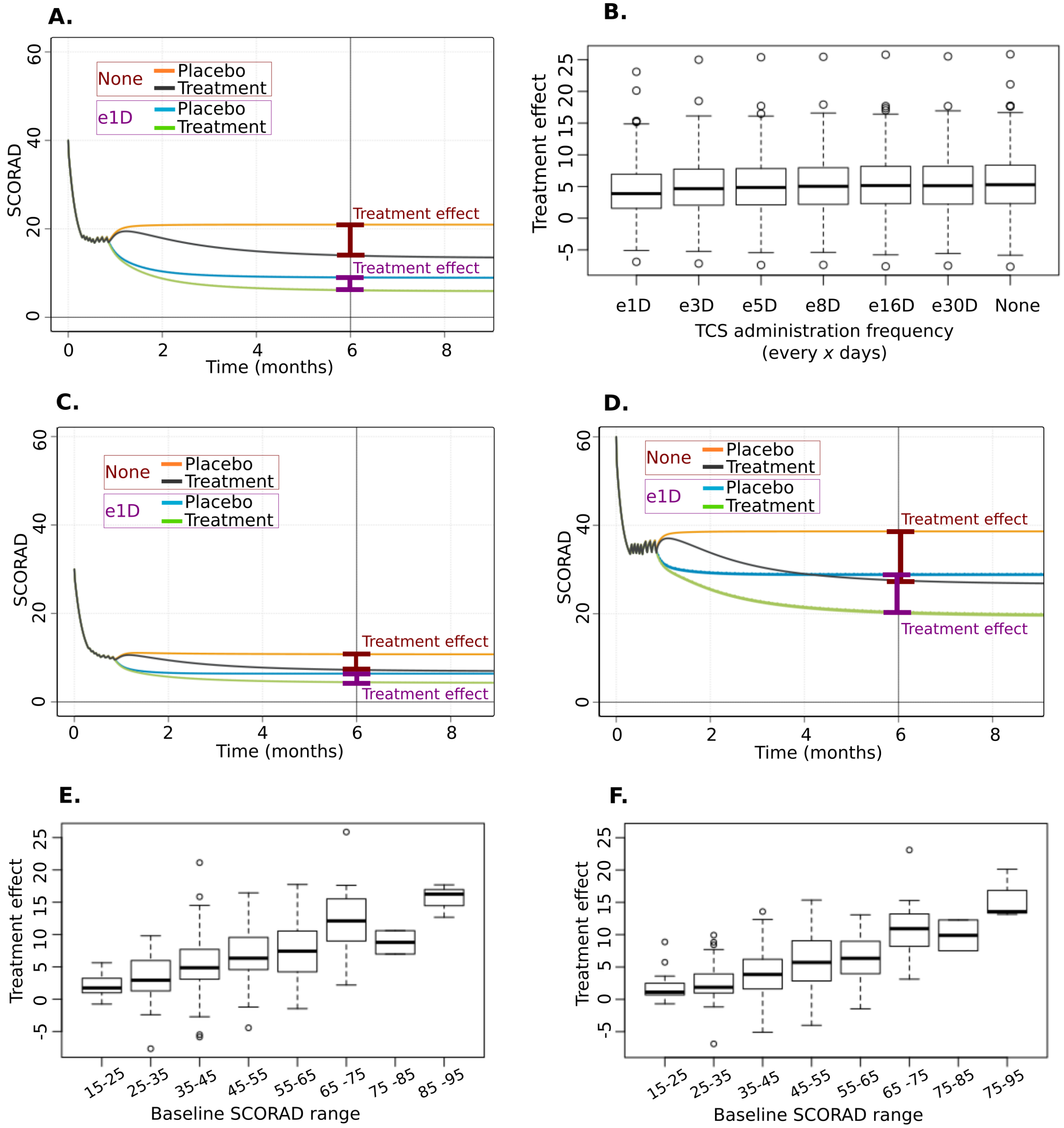

**Figure S8.** Evaluation of the goodness of patient response prediction and the potential for clinical trial optimization by the baseline SCORAD only. **A.** Assessment of patient characteristics predictive of OM-85 treatment effect. **B.** Predictivness of patient characteristics depending of baseline SCORAD. **C.** Optimization of a realistic clinical trial by selecting predicted responders based on their baseline SCORAD only: C1 Average treatment effect, C2. Sample size, C3. Recruitment effort.

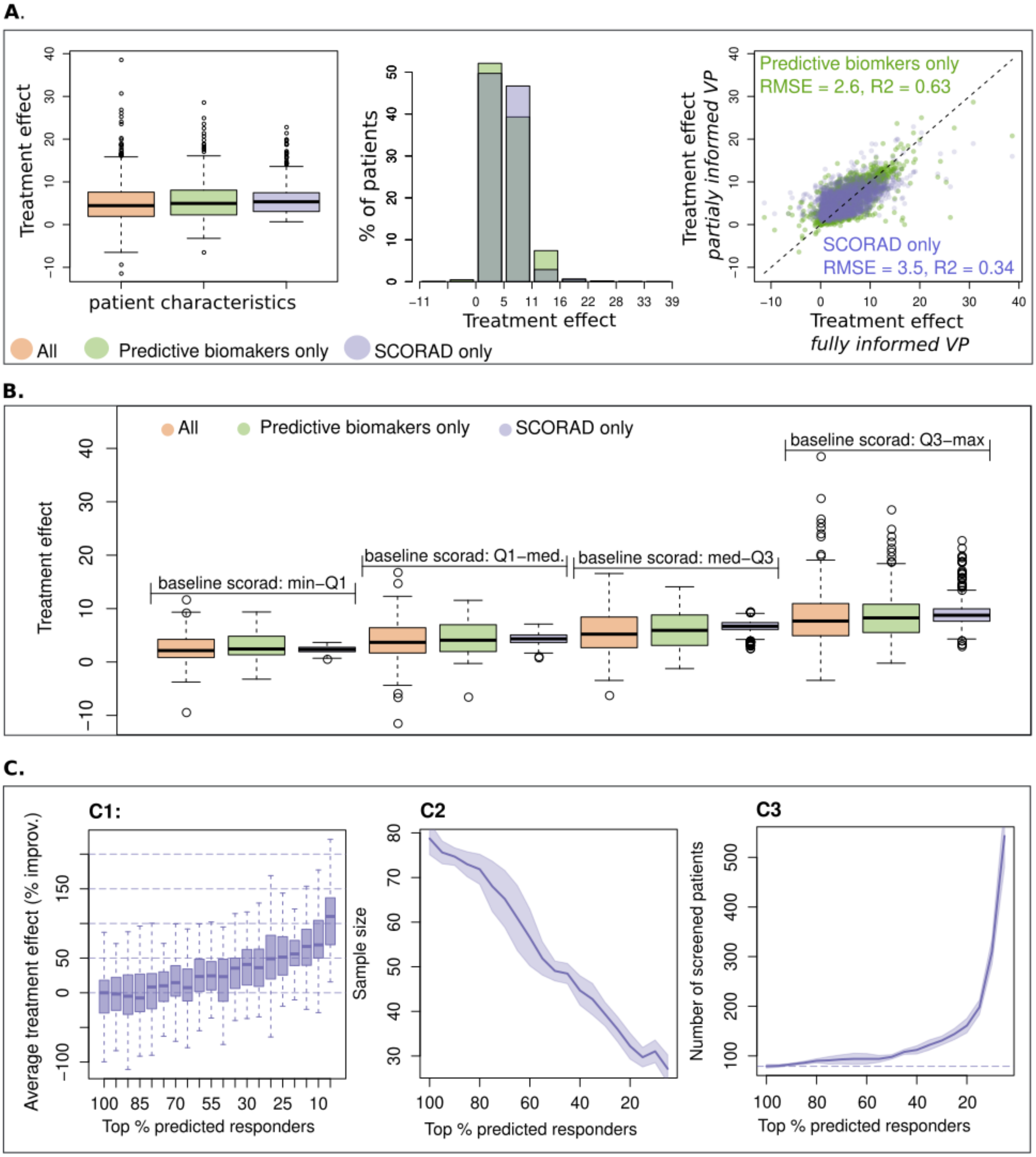

**Figure S9.** Use of a statistical analysis method to identify a set of biomakers predictive of the treatment effect: linear model selection.

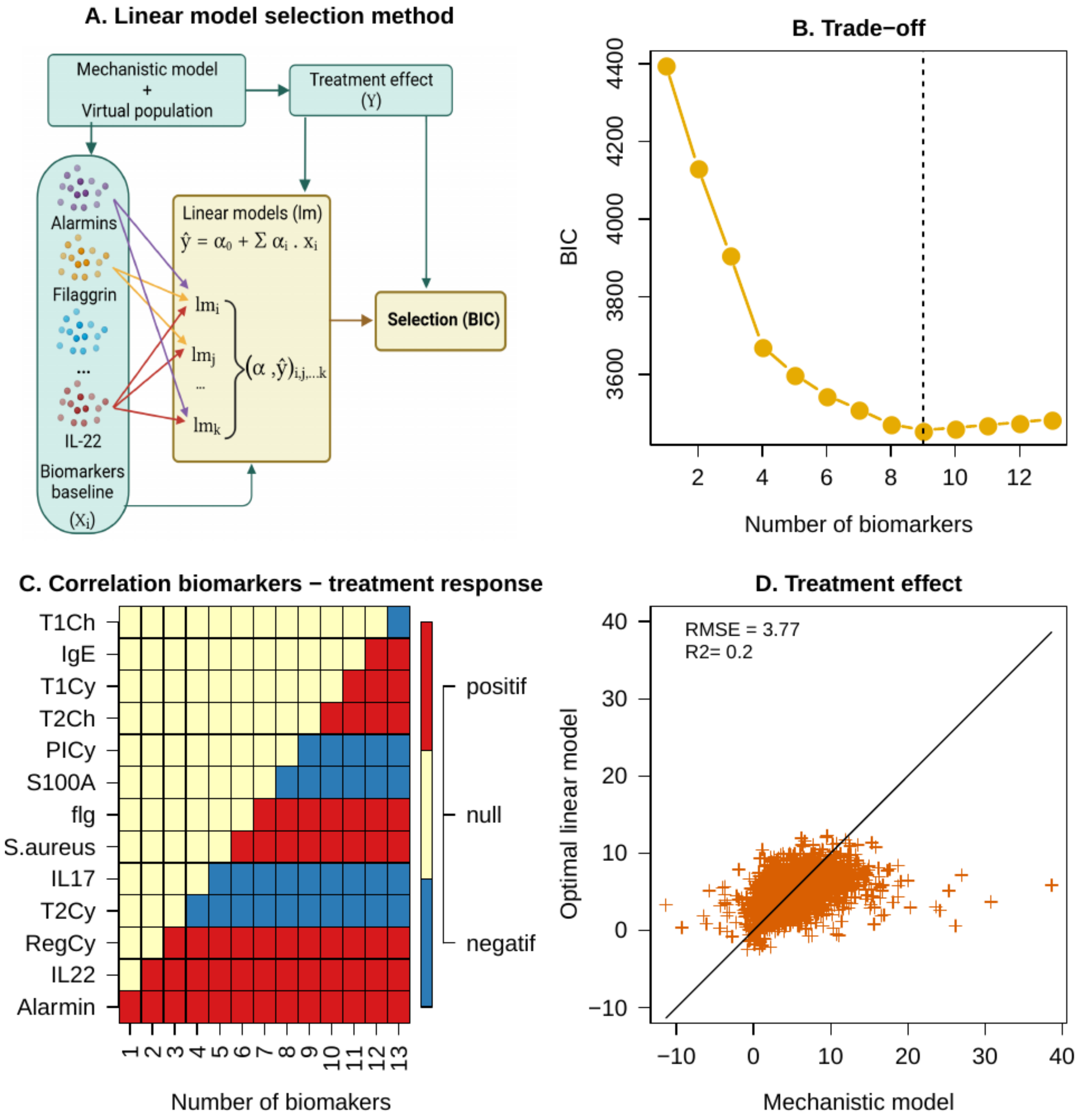

**Figure S10.** Evaluation of the potential for clinical trial optimization by a reduced set of biomarker from the optimal linear model (surrogate model approach).

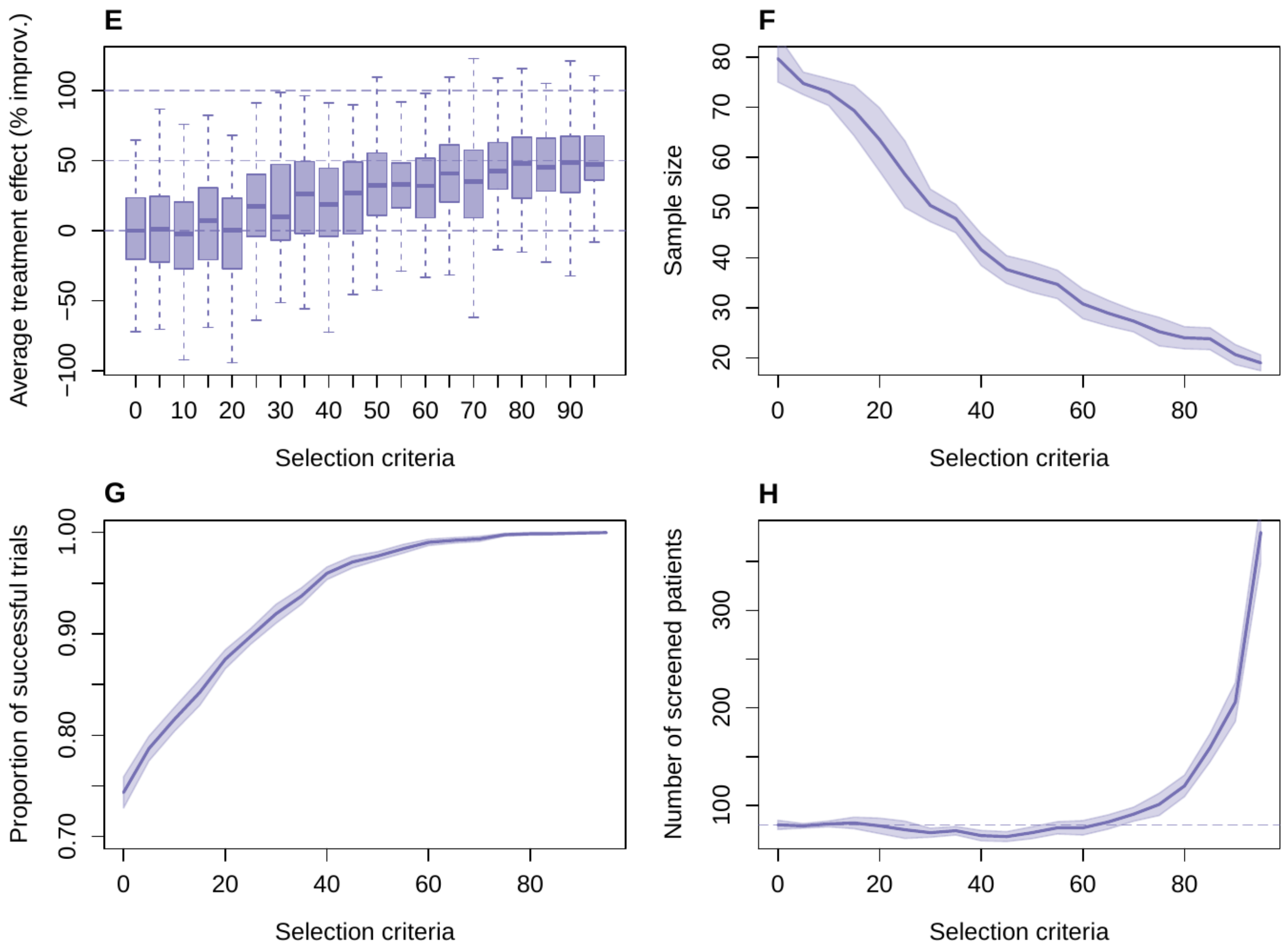

**Figure S11.** Graphical representation of the dimensionality reduction approach

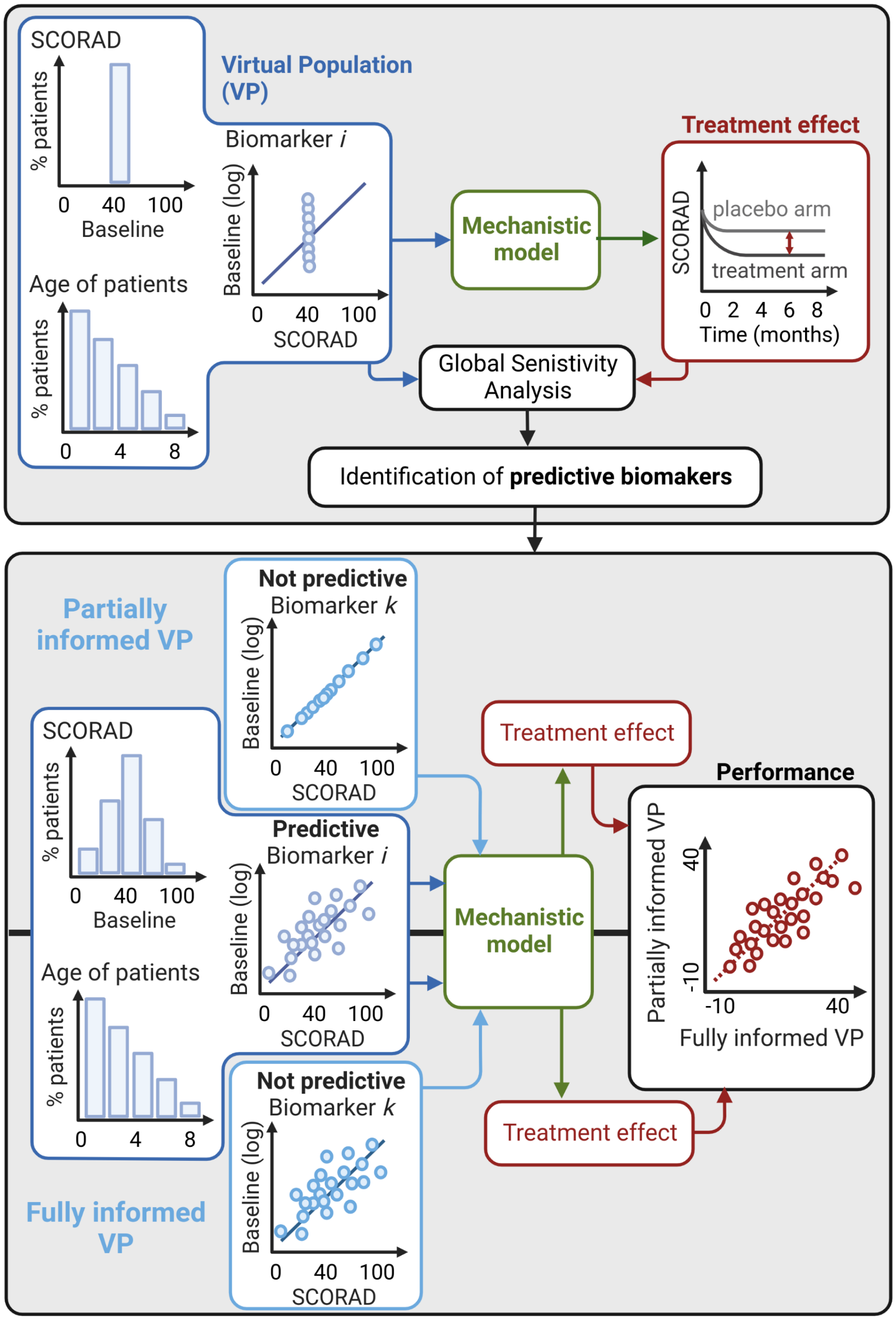

**Figure S12.** Additional results on the identification of predictive biomarkers. **A. Sensitivity indices** (%) from the Global Sensitivity Analysis (GSA): main and between-parameters interactions higher than 0.5 %. **B. Principal Component Analysis** (PCA): predictive biomarkers (arrows) and treatment effect (points). Top responders (blue: worst to red: best) are overall associated with high levels of alarmins and regulatory cytokines (characterizing the first dimension) while low levels of IL-22 and type-2 cytokines (characterizing the second dimension).

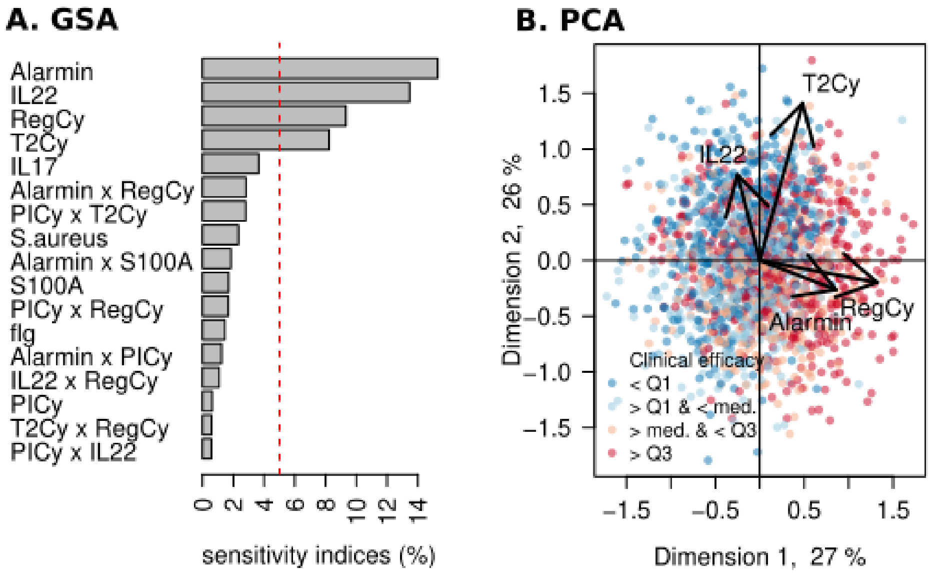

#### S4 Supplementary Tables

**Table S1.** List of biological entities (*i.e.* model variables) represented in the Atopic Dermatitis disease model including under treatment, and corresponding equations. † indicates the biomarkers.

| Entity | | Description | Eq. |
|---|---|---|---|
| *AD severity* | | | |
| EASI | | Eczema Area and Severity Index | (S23) |
| SCORAD | | SCORing Atopic Dermatitis | (S24) |
| *Skin barrier* | | | |
| Skin barrier integrity | $S$ | Qualitative level of the skin barrier integrity | (S1) |
| Filaggrin† | $F$ | Level of filaggrin protein | (S2) |
| Keratinocytes | $K$ | Number of keratinocytes cells | (S5) |
| *Evironmental stressors* | | | |
| Pathogens | $P_I$ | Level of pathogen infiltration in the skin tissue | (S4) |
| *S. aureus*† | $P_S$ | % of colonization of the skin microbiome by *S. aureus* | (S3) |
| *Immune cells* | | | |
| ILC2 | $L$ | Levels of type-2 innate lymphocyte cells | (S6) |
| Activated LCs | $D_L$ | Levels of mature and activated Langerhans cells | (S7) |
| Activated IDEC | $D_E$ | Levels of mature and activated Inflammatory Dendritic Epidermal Cells (IDECs) | (S7) |
| Activated IML | $D_M$ | Levels of innate memory like cells pretuned as type-1 dendritic cells (IML) | (S8) |
| Regulatory T-cells | $T_R$ | Levels of regulatory T-cells | (S20) |
| $IgA^+$ plasmablasts | $P_{A_A}$ | Levels of $IgA^+$ plasmablasts | (S22) |
| *Chemokines* | | | |
| Type-1 chemokines† | $H_1$ | Levels of type-1 chemokines represented by CXCL-10 | (S10) |
| Type-2 chemokines† | $H_2$ | Levels of type-2 chemokines, grouping CCL-17, CCL-22, and CCL-17 | (S11) |
| *Cytokines* | | | |
| Alarmins† | $Y_A$ | Levels of alarmin signals, grouping TSLP, IL-33, and IL-25 | (S13) |
| Pro-Infl. cytokines† | $Y_I$ | Levels of pro-inflammatory cytokines, grouping IL-1$\beta$, IL-6, TNF-$\alpha$, and IL-8 | (S14) |
| Type 1 cytokines† | $Y_1$ | Levels of type-1 cytokines, grouping IL-12 and IFN-$\gamma$ | (S15) |
| Type 2 cytokines† | $Y_2$ | Levels of type-2 cytokines, grouping IL-4 and IL-13 | (S16) |
| IL-17 cytokine† | $Y_{17}$ | Levels of IL-17 cytokine | (S17) |
| IL-22 cytokine† | $Y_{22}$ | Levels of IL-22 cytokine | (S18) |
| S100A proteins† | $Y_S$ | Levels of anti-microbial proteins S100A | (S19) |
| Regulatory cytokines† | $Y_R$ | Levels of regulatory cytokines, grouping IL-10 and TGF-$\beta$ | (S20) |
| *Antibodies* | | | |
| Classe E antibody† | $A_E$ | Levels of polyclonal IgE | (S21) |
| Classe A antibody | $A_A$ | Levels of polyclonal IgA | (S22) |
| *SoC treatments* | | | |
| Emollients | $E$ | Emollient application (binary) | – |
| TCS | $C$ | Topical CorticoSteroids (TCS) application (binary) | – |

**Table S2.** Model parameter notations in the ODE system. All model parameters are positive, the unit is determined by the formalism of the corresponding mechanism, but no unit is precised when relevant. Most of the model parameters are constants, and in this case, this is precised.

| Notation | Description |
|---|---|
| $\rho_X$ | Release rate (constant) of the immune protein (chemokines, cytokines, antibodies) $X$ |
| $\alpha_X$ | Activation rate (constant) of the immune cell $X$ |
| $r_X$ | Renewal / production rate (constant) of the biological entity $X$ |
| $\mu_X$ | Degradation or the death rate (constant) of the biological entity $X$ |
| $c_X$ | Population carrying capacity (constant) of the biological entity $X$ |
| $s_{X1}^{X2}$ | Scaling factor (constant, no unit) linking the biological entity $X1$ to $X2$ |
| $\mathcal{R}_{+/-}^{X}$ | Regulations on mechanism formalized as Hill functions of the modulator concentration $X$ (Tab. S3)<br>$\mathcal{R}_{+}$: up-regulation (activation of amplification)<br>$\mathcal{R}_{-}$: down-regulation |
| $v_{max}^{X}$ | Saturation constant (or maximum reaction rate) of the Hill function (constant) |
| $k_M^X$ | Half-saturation constant of the Hill function (constant) |
| $\mathcal{R}_{env}^{X}$ | Cytokine environment regulating the release of the immune protein $X$ (Tab. S3), no unit |
| $p$ | Potency of TCS (mild, medium, or hight) |
| $\delta_X$ | Switch parameter that turns on or off application of the topical treatment $X$ |

**Table S3.** List of regulation (see Box S1 for the formalism) included in the model, along with the modulators (By), the biological entities regulated (Of), the main literature references (†: assumed) on which the regulations are based, and the equations where the regulations appears.

| Regulation | By | Of | Ref. | Eq. |
|---|---|---|---|---|
| $\mathcal{R}_{+}^{S,Y}$ | $Y_{22}$, $Y_R$ | $S$ | 17, 18 | (S1) |
| $\mathcal{R}_{-}^{S,P_S}$ | $P_S$ | | 1, 3, 5 | |
| $\mathcal{R}_{-}^{S,Y}$ | $Y_2$, $Y_I$, $Y_S$ | | 26–29 | |
| $\mathcal{R}_{+}^{E,S}$ | $E$ | | 20–25 | |
| $\mathcal{R}_{-}^{F}$ | $Y_2$, $Y_{17}$, $Y_{22}$ | $F$ | 6, 18, 27 | (S2) |
| $\mathcal{R}_{+}^{P_I,Y}$ | $Y_2$ | $P_S$, $P_I$ | 2, 27 | (S3) , (S4) |
| $\mathcal{R}_{-}^{P_I,Y}$ | $Y_S$, $Y_I$, $Y_1$ | | 27 | |
| $\mathcal{R}_{+}^{K}$ | $Y_I$, $Y_2$ | $Y_A$, $H_2$ | 59, 61, 62 | (S13) (S11) |
| $\mathcal{R}_{+}^{Y_A}$ | $Y_A$ | $L$, $D_L$ | 41, 42, 44 | (S6), (S7) |
| $\mathcal{R}_{+}^{D_E}$ | $Y_I$ | $D_E$ | | (S7) |
| $\mathcal{R}_{-}^{Y_R}$ | $Y_R$ | $H_2$, $Y_2$, $H_1$, $Y_1$ | 2, 18, 54, 60 | (S10), (S11),(S15),(S16) |
| $\mathcal{R}_{+}^{\rho D}$ | $Y_A$, $A_E$, $P_I$, $C$ | $Y_I$,$H_2$, $Y_2$, $A_E$, $H_1$, $Y_1$, $Y_{17}$, $Y_{22}$ | 43, 45, 47–52 | (S14), (S10), (S11),(S16), (S21), (S15), (S17), (S18) |
| $\mathcal{R}_{env}^{H_2}$ | $Y_2$, $Y_1$ | $H_2$ | 61 | (S11) |
| $\mathcal{R}_{+}^{Y_2,H_2}$ | $H_2$ | $Y_2$ | 66, 71 | (S16) |
| $\mathcal{R}_{env}^{A_E}$ | $Y_2$, $Y_1$, $Y_R$ | $A_E$ | 84, 85 | (S21) |
| $\mathcal{R}_{env}^{H_1}$ | $Y_2$, $Y_1$ | $H_1$ | † | (S10) |
| $\mathcal{R}_{+}^{Y_1,H_1}$ | $H_1$ | $Y_1$ | 66, 69 | (S15) |
| $\mathcal{R}_{+}^{T_{17}}$ | $Y_R$, $Y_I$ | $Y_{17}$, $Y_{22}$ | 42, 46, 72 | (S17), (S18) |
| $\mathcal{R}_{-}^{Y_{17}}$ | $Y_1$, $Y_2$ | $Y_{17}$ | 73 | (S17) |
| $\mathcal{R}_{+}^{Y_{22}}$ | $Y_I$ | $Y_{22}$ | 72 | (S18) |
| $\mathcal{R}_{+}^{Y_S,Y_{22}}$ | $Y_{22}$ | $Y_S$ | 17, 18, 27 | (S19) |
| $\mathcal{R}_{+}^{Y_S,Y_{17}}$ | $Y_{17}$ | | | |
| $\mathcal{R}_{+}^{C,\mu D}$ | $C$ | $D_L$, $D_E$ | 47–52 | (S7) |
| $\mathcal{R}_{-}^{C,infl}$ | | $Y_I$, $Y_1$, $Y_2$ | 47, 50–52, 67, 68 | (S14), (S15), (S16) |
| $\mathcal{R}_{+}^{C,Y_R}$ | | $Y_R$ | 50, 52, 68, 70, 74–77 | (S20) |

**Table S4.** Estimated relationships between baseline biomarkers and AD severity. Notations: for a given $X$ biomarker, the baseline value is $X_{Bas}$ (note that $X_{Bas} = (X)^{\star}$); reference (*i.e.* average) value depending on AD severity is denoted $X_{Bas}^{Ref}$; the white noise *i.e.* the between-patient variability distributed around the average value is denoted $\sigma_X$. References of literature data on which the estimates are based are provided.

| Biomarker | At reference | Between-patient variability | Ref |
|---|---|---|---|
| $P_S$ | $P_{S\,Bas}^{Ref} = 1 + \frac{99 \times (SCORAD_{Bas})^2}{74^2 + (SCORAD_{Bas})^2}$ | $P_{S\,Bas} = \frac{\sigma_{PS} \times P_{S\,Bas}^{Ref}}{1 + P_{S\,Bas}^{Ref} \times (\sigma_{PS} - 1)}$ <br> $\sigma_{PS} \sim Lognormal(0, 1)$ | 106, 107 |
| $F$ | $F_{Bas}^{Ref} = e^{-0.02 \times SCORAD_{Bas} + 4.5}$ | $F_{Bas} = F_{Bas}^{Ref} \times e^{\sigma_F}$ <br> $\sigma_F = \mathcal{N}(0, 0.125)$ | 105 |
| $Bmk$ | $Bmk_{Bas}^{Ref} = e^{\alpha_{Bmk} \times SCORAD + log(Bmk_{HC})}$ | $Bmk_{Bas} = Bmk_{Bas}^{Ref} \times e^{\sigma_{Bmk}}$ <br> $\sigma_{Bmk} \sim \mathcal{N}(0, 1.25)$ | 101, 103, 104 |
| $Y_I$ | $\{\alpha_{YI} = 0.0675\ ,\ Y_{IHC} = 67.1\}$ | | |
| $H_2$ | $\{\alpha_{H2} = 0.0538\ ,\ H_{2HC} = 115\}$ | | |
| $Y_S$ | $\{\alpha_{YS} = 0.111\ ,\ Y_{SHC} = 8.7\}$ | | |
| $Low$ | $\{\alpha_{Low} = 0.005\ ,\ Low_{HC} = 0\}$ | for $Low := \{Y_A, Y_2, H_1, Y_1, Y_{17}, Y_{22}, Y_R, A_E\}$ | |

**Table S5.** Summarized levels (in pg/mL) of AD severity and immune biomarkers in lesional skin of atopic dermatitis children (AD) and in healthy skin of control children (HC), directly extracted or extrapolated from Lyubchenko *et al.* (2020)[101] and as used for the model calibration. $^{\dagger}$ corresponds to biomarkers with levels assumed below the LoQ = 50 pg/mL based on Lyubchenko *et al.* (2020) and / or He *et al.* (2020) [103, Fig.2, 3, 5] and Guttman-Yassky *et al.* (2019) [104, Fig. 1B, 2A]. Biomarker expression (RT-PCR quantification from skin tape strips) from He *et al.* (2020) and Guttman-Yassky *et al.* (2019) are also used to complement missing data from Lyubchenko et al. (2021). Reported values are the minimal min, first quartile Q-1, median med., third quartile Q-3, and maximal max concentrations and are extracted from the individual absolute concentrations in pg/mL.

| *SCORAD* | AD | 16.8 | 32.3 | 40.4 | 54.2 | 78.2 | |
|---|---|---|---|---|---|---|---|
| Biomarker | Patient | Min | Q-1 | Med. | Q-3 | Max | AD vs HC |
| $Y_A^{\dagger}$[101] | AD | | | | | 50 | AD $\approx$ HC[104,110]; |
| | HC | | | 0 | | | AD $>$ HC[110] |
| $Y_I$[101] | AD | 65.1 | 318 | 966 | 3.94e3 | 2.35e4 | AD $\gg$ HC[101,104] |
| | HC | 7.9 | 27.7 | 67.1 | 112 | 1.09e3 | |
| $Y_2^{\dagger}$[101,104] | AD | | | | | 50 | AD $\gg$ HC[104,110] |
| | HC | | | 0 | | | |
| $H_2$[101] | AD | 34.6 | 444 | 1.12e3 | 2.44e3 | 1.97e4 | AD $\gg$ HC |
| | HC | 21.4 | 55.1 | 115 | 431 | 1.28e3 | |
| $Y_1^{\dagger}$[101,103,104] | AD | | | | | 50 | AD $\gg$ HC[104]; |
| | HC | | | 0 | | | AD $\approx$ HC[110] |
| $H_1^{\dagger}$[101,103,104] | AD | | | | | 50 | AD $\approx$ HC[104,110] |
| | HC | | | 0 | | | |
| $Y_{17}^{\dagger}$[101] | AD | | | | | 50 | AD $\gg$ HC[110] |
| | HC | | | 0 | | | |
| $Y_{22}^{\dagger}$[101] | AD | | | | | 50 | AD $\gg$ HC[110] |
| | HC | | | 0 | | | |
| $Y_S$[101,103,104] | AD | 37.7 | 267 | 844 | 3.64e3 | 2.22e4 | AD $\gg$ HC[110] |
| | HC | 0.6 | 4.67 | 8.72 | 32 | 511 | |
| $Y_R^{\dagger}$[101] | AD | | | | | 50 | AD $\gg$ HC[111] |
| | HC | | | 0 | | | |
| $A_E^{\dagger}$ | AD | | | | | 50 | AD $\gg$ HC |
| | HC | | | 0 | | | |

**Table S6.** Summary of data used for the calibration of patient response to standard of care. All datasets have been homogenized to use the median, narrow and wide bounds of the relative improvement percentage of EASI

| Reference | Treatment | Cohort | Data used |
|---|---|---|---|
| Patrizi et al (2008)[112] | Emollients | 2-13 y.o, 20 AD (mild to moderate) | Time series, at 7, 14, 21, 28 and 42 days |
| Szczepanowska et al. (2008)[113] | Emollient + TCS, TCS alone | 2-15 y.o., 26 AD (mild to moderate) | Time series, at 7 and 14 days |
| Kim et al. (2020)[114] | Mild TCS | Adults, 52 AD (mild to moderate) | Time series, at 14 and 28 days |
| Luger et al. (2004)[115] | Emollient + mild TCS | Adults, 300 AD (moderate to severe) | Time series, at 8, 22,43,90 and 120 days |
| Miyano et al. (2022)[87] | Emollient + TCS | Aggregated from several studies (moderate to severe), baseline EASI = 25 | Time point at 14 days |

**Table S7.** Calibrated parameter values by fitting the reference patient to SoC calibration data. NU: no units

| Name | Unit | Calibrated value |
|---|---|---|
| $k_S^E$ | 1/day | 3.1973 |
| $k_{\mu D}^C$ | NU | 1.206 |
| $k_{YR}^C$ | NU | 7.44 |
| $k_{\rho D}^C$ | NU | 0.414 |
| $k_{infl}^C$ | NU | 0.142 |
| $s_p$ | NU | 0.217 |
| $k_{MAct,Y_A}$ | pg/mL | 2.265 |
| $s_{DE}$ | NU | 2146.87 |
| $v_{maxD_L}$ | NU | 1.5157e-5 |
| $\mu_{PI}$ | 1/day | 0.1687 |
| $v_{maxS,}$ | 1/day | 0.1 |
| $v_{maxS,Y}$ | 1/day | 2.92 |
| $v_{maxS,Y}$ | NU | 16.04 |
| $v_{maxK}$ | NU | 1.896 |
| $v_{maxY_S,Y_{17}}$ | NU | 17.671 |
| $v_{maxY_S,Y_{22}}$ | NU | 13.01 |
| $v_{maxS,Y_2}$ | NU | 0.198 |
| $k_{MS,Y_I}$ | NU | 0.4324 |
| $v_{maxS,Y_2 \times Y_I}$ | NU | 1.01 |
| $v_{maxS,Y_S}$ | NU | 1.06 |
| $s_{Y_2,D_L}$ | NU | 173.05 |
| $s_{D_L,nD_L}$ | NU | 1879.76 |
| $\alpha_L$ | cells/(mL x day) | 2.5468e4 |
